# Genetic variant in *SPDL1* reveals novel mechanism linking pulmonary fibrosis risk and cancer protection

**DOI:** 10.1101/2021.05.07.21255988

**Authors:** Jukka T. Koskela, Paavo Häppölä, Aoxing Liu, FinnGen, Juulia Partanen, Giulio Genovese, Mykyta Artomov, Mikko N.M. Myllymäki, Masahiro Kanai, Wei Zhou, Juha M. Karjalainen, Teemu Palviainen, Justiina Ronkainen, Sylvain Sebert, Taru Tukiainen, Priit Palta, Jaakko Kaprio, Mitja Kurki, Andrea Ganna, Aarno Palotie, Tarja Laitinen, Marjukka Myllärniemi, Mark J. Daly

**Affiliations:** Institute for Molecular Medicine Finland (FIMM), University of Helsinki, Helsinki, Finland; Medical and Population Genetics, Broad Institute, Cambridge, MA, United States of America; Stanley Center, Broad Institute of Harvard and MIT, Cambridge, MA, United States of America; Hematology Research Unit Helsinki, Helsinki University Hospital Comprehensive Cancer Center, 00290, Helsinki, Finland; Department of Computational Medicine and Bioinformatics, University of Michigan, Ann Arbor, MI, USA; Center for Statistical Genetics, University of Michigan School of Public Health, Ann Arbor, MI, USA; Analytical and Translational Genetics Unit, Massachusetts General Hospital, Boston, MA, USA; Center for Life Course Health Research, University of Oulu, Oulu, Finland; Administration Center, Tampere University Hospital, Tampere, Finland; University of Helsinki and Helsinki University Hospital, Heart and Lung Center, Department of Pulmonary Medicine, Helsinki, Finland

## Abstract

Idiopathic Pulmonary Fibrosis (IPF) is a rare disease with poor prognosis. By contrast, cancer is common in any elderly population and a leading killer, but is now often curable. Of note, whereas IPF is driven by cellular senescence, cancer is characterized by uncontrolled cell division.

Using data available from two large biobank-based studies (Finnish FinnGen study and UK biobank), we conducted a comprehensive analysis of the shared genetic background of IPF and cancer. In a population sample of 218,792 Finns with complete longitudinal health histories, we estimated the effect of individual genetic variants to the lifetime risk of IPF and cancer. We extend the analysis from IPF-GWAS to pan-cancer meta-analysis over FinnGen and UK Biobank and finally to the identification of genetic drivers of somatic chromosomal alterations.

We detected six loci (*SPDL1, MAD1L1, MAP2K1, RTEL1-STMN3, TERC-ACTRT3, OBFC1*) associated with both IPF and cancer, all closely related to cellular division. However, each individual signal is found with opposite effects over the two diseases, termed as antagonistic pleiotropy. Several of these loci (*TERC-ACTRT3, RTEL1-STMN3, OBFC1*) are among the strongest inherited factors for constitutive telomere length variation and consistently indicate that shorter telomere length would increase the risk for IPF but protect from malignancy. However, a Finnish enriched *SPDL1* missense variant and a common *MAD1L1* intronic variant had no effect on telomere length but were shown to protect individuals from accumulation of somatic mutations.

The decreased risk of cancer in *SPDL1* and *MAD1L1* variant carriers might result from a lower number of chromosomal alterations accumulated over time, conversely leading to fibrosis in the lung due to cellular senescence-induced inflammation. We hypothesize that the *SPDL1* missense variant functions as gain-of-function mutation, leading to cellular senescence, a barrier to cancer and a driver of fibrosis in IPF. If translated to therapy, these findings might not only be able to offer relief to individuals with IPF, but also to protect from onset of cancer.

## 1 INTRODUCTION

Idiopathic Pulmonary Fibrosis (IPF) is the most common Idiopathic Interstitial Pneumonias (IIPs) affecting 14-63/100,000 individuals with a typical onset around 60-70 years of age,^1^ characterized by progressive and diffuse fibrosis of the lung. The prognosis of the condition is poor with a mean post-diagnostic survival until death or lung transplantation of 3-4 years. ^2^ Two pharmacotherapeutic options exist and have been shown to increase life-expectancy,^3,4^ but offer no symptoms relief. ^5,6^

IPF presents both in sporadic and familial forms,^7,8^ while the latter is thought to represent 3-15% of all IPF. The most recent Genome-wide Association Study (GWAS)identified three new loci,^9^ currently resulting in 20 risk loci not all replicating over studies.

IPF, together with aplastic anemia and liver cirrhosis, are typical manifestations of telomere biology disorders presenting in adulthood,^10^ while mutations of few key telomere associated genes are thought to account for up to 15-25% of familial and less than half of that in sporadic IPF.^11,12^ Telomeres shorten in every cell division during normal aging finally causing telomere exhaustion and replicative senescence,^13^ a driver of fibrotic changes in the lung,^14^ also independently of telomere attrition.^15^

Telomere attrition, genomic instability and cellular senescence are among the hallmarks of aging,^16^ while aging is the most significant risk factor to cancer – a second leading cause of death globally.^17^ Importantly however, cellular senescence and apoptosis act as primary tumor suppressors,^18,19^ but are the drivers of fibrosis in IPF ^20^ through growth arrest and Senescence-Associated Secretory Phenotype.^21,22,23^

In this study we genetically confirm a mechanistic link between IPF and cancer, connected by a single inherited variant, highly enriched in Finnish population. Dramatically different in nature, the specific genetic variant is shown to increase IPF risk while decreasing risk of common forms of cancer – a prime example of antagonistic pleiotropy.^24^ With this observation in mind, we proceed to identify a number of other convincing genetic links connecting the two in the same way, which reveal additional mechanisms relevant to both diseases, all generally related to cell division.

## 2 MATERIAL AND METHODS

### 2.1 Study participants and disease endpoints

The FinnGen (in preparation) study derives disease phenotypes from nationwide health registries - including the national cancer registry dating back to 1953, and national hospital discharge and causes of death registries established in 1969 which together include all neoplasms, inpatient hospital visits, and deaths in Finland. These registries, and therefore phenotypes of individuals in FinnGen, include all events since birth (or initiation of the registry) regardless of the age of enrollment to the study. Loss to follow-up can occur only due to moving abroad. Definition of diseases is discussed in the Supplementary Appendix. Data release V & VI were used in the present study, while the most recent public Data release IV of FinnGen included 176,899 individuals and 2,264 phenotypes, available at http://r4.finngen.fi/.

In addition to recruiting participants from Finnish biobanks, FinnGen has focused recruitments, such as from the national IPF registry project FinnishIPF, ^25^ which includes 873 patients meeting ATS/ERS criteria of IPF,^26^ of which 240 patients have been included in FinnGen.

Replication of specific findings was done in the UK biobank (UKB),^27^ Northern Finnish Birth Cohort 1966 (NFBC1966),^28^ and Finnish Twin Cohort (FTC).^29^

### 2.4 Statistical analysis

Genome-wide analysis of each disease endpoint in FinnGen was conducted with SAIGE ^30^ and adjusted for age, sex, genotyping batch and platform together with ten first principal components. Fine-mapping of genetic loci in FinnGen was performed using SuSiE.^31^ A credible set of varying numbers of markers is identified, in an aim to capture each independent signal in a separate credible set while each variant in a specific credible set is ranked by the posterior probability of being causal. The variant with highest probability of being causal is discussed in results. All effects are reported for the GRCh38 alternative allele.

To account for competing risks, cumulative incidence functions were based on Aalen-Johansen estimates and survival analyses were conducted using cause-specific Cox regression models. Cox models were stratified by sex to allow different baseline hazards for men and women and adjusted with first two genetic principal components and their non-linear effects using penalized smoothing splines. Covariates demonstrating non-proportional hazards were treated as time-dependent.

To investigate the relationship between inherited genetic variants and mosaic Chromosomal Alteration (mCA) events in FinnGen participants, a logistic regression model was used. We regressed the status of carrying mCA event on the allele dosage of the variant under assesment, adjusting for age, age^2^, sex, and ten first principal components in addition to smoking status.

### 2.5 Calling somatic variants from array intensity data

Intensity data from genotyping was transformed into log2R ratio (LRR) and B-allele frequency (BAF) values and used to determine the local imbalances in allelic fractions.^32^

### 2.6 Analytical strategy

We first explored novel genetic associations identified in the FinnGen IPF GWAS and their cross-replication in the largest IPF meta-analysis to date.^9^ We then related these findings to the telomere function and explored their associations to other diseases and health related conditions defined in FinnGen (n=2,925). After observing a notable pleiotropy with common cancers, we used longitudinal registries to investigate the age of onset considering also the competing risks of other diseases of interest. Finally, we describe the function of the novel linking element and show that the pathology of these diseases is connected (but with opposite genetic effects) to multiple other genetic loci, many related to cell division.

## 3 RESULTS

The FinnGen study (Data release V) consists of 218,792 participants including 1,028 patients with IPF and 38,036 patients with cancer, Table 1.

**Table 1.** Finngen participants with IPF or cancer at the end of the study follow-up. Male breast cancer excluded.

| Disease | Cases (%) | Women (%) | Median onset age (IQR) | Cause-specific deaths (%) | Median years to cause-specific death (IQR) |
| --- | --- | --- | --- | --- | --- |
| Colorectal cancer | 3,022 (1.4%) | 1,315 (43.5%) | 66.7 (58.4 - 73.8) | 489 (16.2%) | 1.2 (0.3 - 2.8) |
| Prostate cancer | 6,311 (2.9%) | 0 (0.0%) | 68.1 (62.6 - 73.6) | 484 (7.7%) | 5.1 (2.0 - 10.0) |
| Breast cancer | 8,401 (3.8%) | 8,401 (100.0%) | 58.6 (50.4 - 66.3) | 334 (4.0%) | 5.4 (2.2 - 10.7) |
| Nonmelanoma skin cancer | 10,382 (4.7%) | 5,142 (49.5%) | 65.6 (55.7 - 73.3) | 14 (0.1%) | 2.1 (0.4 - 12.9) |
| Any cancer | 38,036 (17.4%) | 21,066 (55.4%) | 62.4 (52.1 - 70.4) | 4,107 (10.8%) | 2.0 (0.4 - 7.1) |
| IPF | 1,028 (0.5%) | 378 (36.8%) | 70.3 (62.3 - 76.8) | 176 (17.1%) | 0.8 (0 - 4.6) |

### 3.1 Genome-wide association study of IPF in FinnGen identifies 3 new Finnish-enriched variants

GWAS of 1,028 IPF patients and 196,986 controls replicated (at P<0·05) 9 out of the 13 independent loci (Table 2) confirmed in the latest meta-analysis,^9^ whereas one variant (rs2077551) was not imputed in FinnGen. Moreover, we identify three new loci (*SPDL1, TACC2* and *AXIN1*, more detailed discussion of the latter two with Manhattan and quantile-quantile plots can be found in the Supplementary appendix) associated with IPF and reaching genome-wide significance (P<5×10^−8^).

**Table 2.** IPF associated variants in FinnGen reaching genome-wide significance (P<5×10^−8^) and replication of previously reported IPF variants from Allen et al. (2020). Enrichment ratio is based on allele frequencies in gnomAD. Effects reported for the GRCh38 alternative allele.

| Variant | Nearest gene | Predicted effect | Ref/Alt | AF<sub>Alt</sub> | Enrichment (FIN / NFEE) | OR<sub>Alt</sub> (95% CI) | P |
| --- | --- | --- | --- | --- | --- | --- | --- |
| Novel |  |  |  |  |  |  |  |
| rs116483731 | SPDL1 | Missense | G/A | 0.030 | 4.92 | 3.13 (2.36–4.15) | 9.97E-16 |
| rs967235139 | TACC2 | Intronic | T/C | 0.0021 | Not seen in NFEE | 35.86 (9.83–130.74) | 4.30E-08 |
| rs185488877 | AXIN1 | Intronic | C/T | 0.019 | 9.52 | 2.74 (1.92–3.92) | 2.32E-08 |
| rs770066110 | TERT | Stop | G/A | 0.00061 | Not seen in NFEE | 9833.99 (700.03–138146.80) | 9.16E-12 |
| rs776981958 | TERT | Missense | T/C | 0.0016 | Not seen in NFEE | 1029.96 (241.74–4388.40) | 6.53E-21 |
| Previously reported in Allen et al. (2020) |  |  |  |  |  |  |  |
| rs78238620 | KIF15 | Intronic | T/A | 0.040 | 0.67 | 1.29 (1.02–1.62) | 3.03E-02 |
| rs2293607 | TERC-ACTRT3 | Non coding transcript exon | T/C | 0.27 | 1.08 | 1.30 (1.17–1.44) | 5.99E-07 |
| rs2013701 | FAM13A | Intronic | G/T | 0.50 | 1.02 | 0.85 (0.78–0.93) | 3.16E-04 |
| rs7725218 | TERT | Intronic | G/A | 0.36 | 1.03 | 0.80 (0.73–0.88) | 3.26E-06 |
| rs2076295 | DSP | Intronic | T/G | 0.38 | 0.82 | 1.24 (1.13–1.36) | 4.96E-06 |
| rs12699415 | MAD1L1 | Intronic | G/A | 0.41 | 1.04 | 1.06 (0.96–1.16) | 2.20E-01 |
| rs2897075 | ZKSCAN1 | Intronic | C/T | 0.39 | 1.05 | 1.17 (1.07–1.29) | 7.26E-04 |
| rs28513081 | DEPTOR | Intronic | A/G | 0.50 | 1.18 | 0.97 (0.89–1.06) | 5.00E-01 |
| rs35705950 | MUC5B | Upstream gene variant | G/T | 0.10 | 0.96 | 4.98 (4.21–5.89) | 3.88E-80 |
| rs9577395 | ATP11A | Intronic | C/G | 0.22 | 1.02 | 0.92 (0.83–1.03) | 1.39E-01 |
| rs59424629 | IVD | Intronic | G/T | 0.61 | 1.16 | 1.11 (1.01–1.22) | 2.34E-02 |
| rs62023891 | AKAP13 | Intronic | G/A | 0.31 | 1.13 | 1.10 (0.99–1.21) | 6.15E-02 |
| rs12610495 | DPP9 | Missense | A/G | 0.34 | 1.15 | 1.10 (1.00–1.22) | 3.80E-02 |

We identified a missense variant in *SPDL1* at 5q35.1 (rs116483731, odds ratio for the A allele, 3·13; 95% CI 2·37 to 4·14; P=9·97×10^−16^), recently reported associated with IPF in FinnGen.^33^ Although this variant (p.Arg20Gln) in *SPDL1* is enriched in the Finnish population (Table 2), it was replicated in the IPF meta-analysis MAF 0·8%, odds ratio for the A allele, 2·40; 95% CI 1·80 to 3·40; P=7·55×10^−7^). In FinnGen, the variant was also associated with the risk of receiving a lung transplant (rs116483731, odds ratio for the A allele, 2·34; 95% CI 1·05 to 5·23; P=0·0373), as 97 participants had received a transplant for any reason.

Fine-mapping the *TERT* locus identified two independent putative functional risk variants at 5p15.33, both located in the reverse transcriptase domain of *TERT*, known to harbor several telomere disease mutations. First, we report an extremely rare Loss-of-Function (LoF) mutation (p.Arg774X, high confidence ^34^ LoF) in *TERT* gene (rs770066110, odds ratio for the A allele, 9833·99; 95% CI 700·03 to 138146·8; P=9·16×10^−12^). Second, we identified a rare missense mutation (p.Asp684Gly) in the *TERT* gene (rs776981958, odds ratio for the C allele, 1029·96; 95% CI 241·74 to 4388·40; P=6·53×10^−21^) also independently associated with IPF. According to the gnomAD resource, both functional *TERT* mutations are only polymorphic in Finnish, Swedish or Estonian populations. The effect on Lymphocyte Telomere Length (LTL) was confirmed in NFBC1966 and FTC cohorts,^28,29^ as discussed in the Supplementary Appendix.

### 3.3 SPDL1 missense variant increases IPF risk but decreases the risk of common cancers

Next, we searched the FinnGen data for any other effects the *SPDL1* missense variant rs116483731 might carry over numerous conditions. Strikingly, the variant was found to confer protection over different types of cancer with a considerable risk reduction for any cancer (odds ratio for the A allele, 0·81; 95% CI 0·77 to 0·85; P=2·05×10^−15^). The only risk increasing effect was seen for ILD, IPF and their complications at P<1×10^−5^ (Figure 1, Supplementary Table S2).

**Figure 1.**
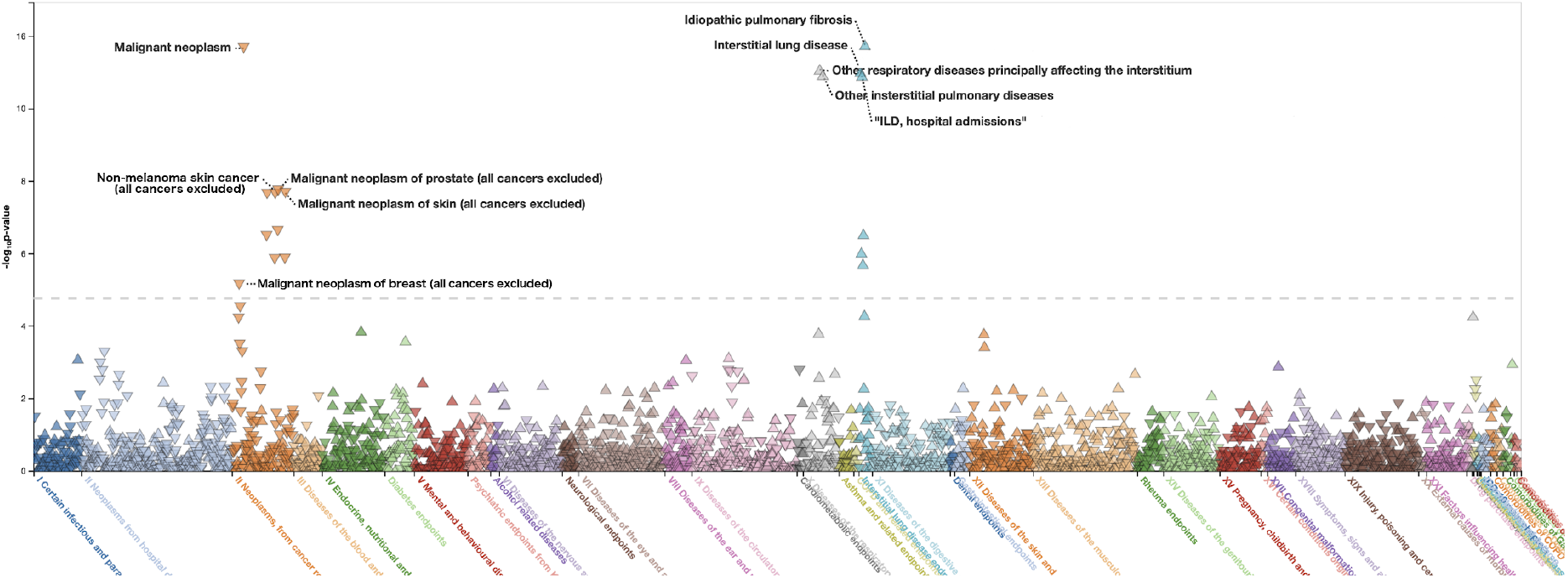
Disease and health related endpoint associations for rs116483731 in a phenome-wide plot in FinnGen, n endpoints=2925. Upward triangle represents increased risk to a disease, downward triangle represents protection from disease.

The expression of *SPDL1* was shown to be increased in 9 out of 11 cancer types in the The Cancer Genome Atlas Research Network (https://www.cancer.gov/tcga), further discussed in the Supplementary Appendix.

We also assessed other functional variants (12 alleles) in the *SPDL1* gene including a loss-of-function variant. None of these functional variants were associated with any cancer or ILD-related endpoints, Table S2.

### 3.4 Cancer incidence over time in SPDL1 carriers

To further assess the observed opposite effects of rs11648373 to lifetime risk of IPF and cancer (and its subtypes) in a competing risk context, we analyzed the FinnGen data with cause-specific Cox proportional hazards models and cumulative incidence estimates. We separated participants by their disease status at study entry, referred to as incident, prevalent and as a combination of both.

The risk of IPF in the carriers was of similar magnitude compared to cross-sectional analysis when assessed longitudinally (hazard ratio 2·27 for the A allele; 95% CI 1·86 to 2·76; P=4·46×10^−16^) combining incident and prevalent cases (Figure 2, Supplementary Table S3). We also confirmed the lower risk of any cancer (hazard ratio 0·83 for the A allele; 95% CI 0·79 to 0·87; P=4·23×10^−^ 15). This protective effect was seen individually across diverse cancers, such as prostate, breast, and non-melanoma skin cancer but not in colorectal cancer (Supplementary Table S3).

**Figure 2.**
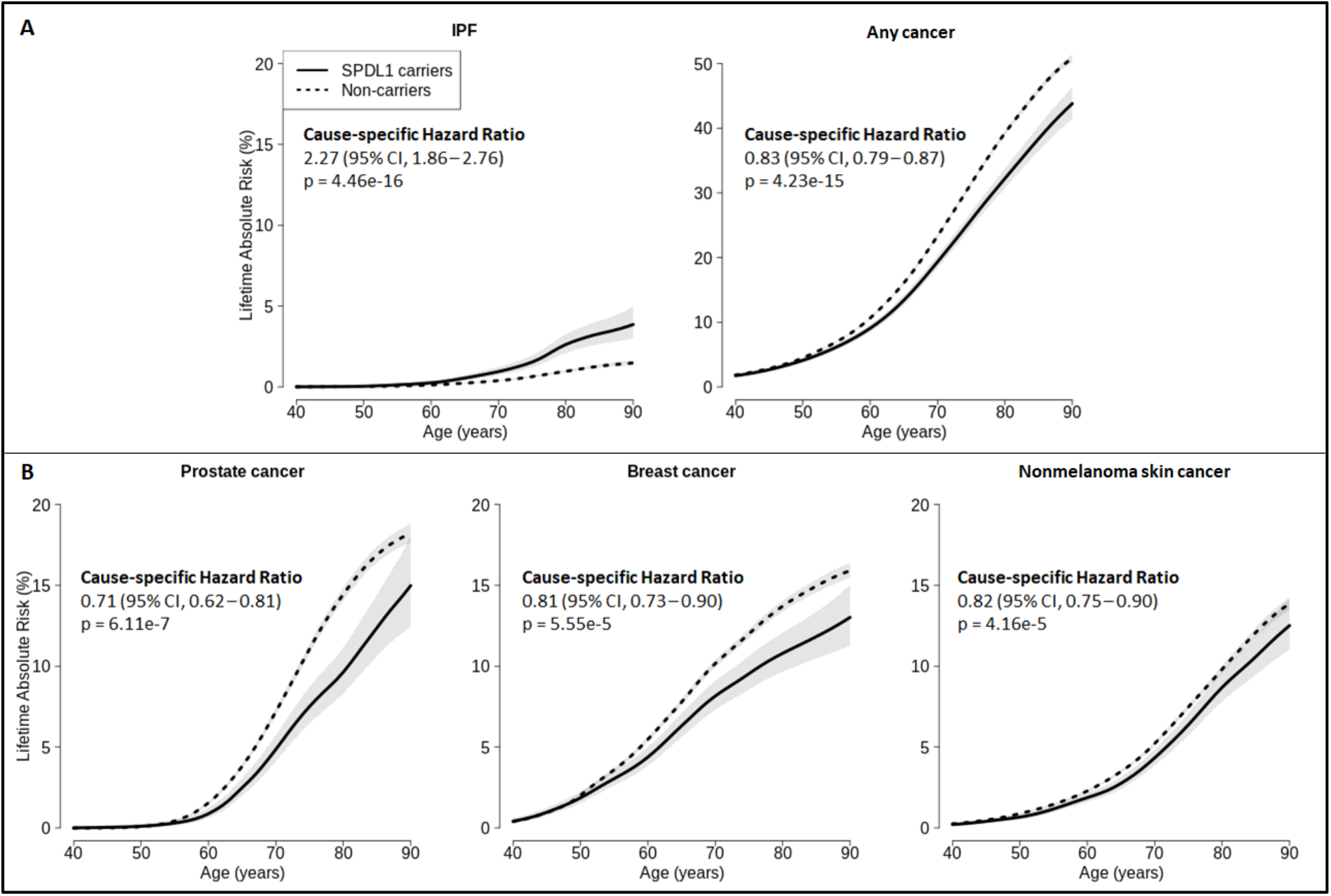
**Cumulative incidence and cause-specific hazard ratios of IPF and cancers in FinnGen, stratified by SPDL1 missense variant rs116483731, considering both prevalent and incident cases at the study entry**.

### 3.7 Further evidences of opposed pleiotropy between IPF and cancer

As antagonistic pleiotropy was observed over IPF and cancer in the case of the *SPDL1* missense variant, we next sought to assess if more such variation could be identified. We performed a pan-cancer meta-analysis in FinnGen and UK Biobank, consisting of 51,271 participants with cancer and 209,134 cancer-free participants in the FinnGen study (Data release VI, not utilized in any other analysis) in addition to 65,783 participants with cancer and 343,346 cancer free participants in UKB, all of White British ancestry. This allowed us to identify causal alleles generally associated with cancer, not solely focusing on any specific cancer type while leveraging power from an increased number of participants.

Pan-cancer meta-analysis over FinnGen and UKB identified 62 loci reaching genome-wide significance (P<5×10^−8^, Supplementary Table S10). Of the 62 loci, 31 were driven by cancer of a specific organ (skin, breast or prostate) and 31 were determined as associated with a broader risk of different cancer types.

We then intersected the 62 lead association signals from the pan-cancer meta-analysis with results from the latest IPF-meta-analysis, resulting in 51 variants found in both studies. Notably, 6 of the 51 variants also had effects (Bonferroni threshold of P<0·0008, Table 3) in the IPF meta-analysis. All of lead variants in the 6 loci (*SPDL1, MAD1L1, MAP2K1, RTEL1-STMN3, TERC-ACTRT3, OBFC1*) presented the same antagonistic pleiotropy over IPF and cancer as *SPDL1*.

**Table 3.** Lead SNVs from pan-cancer analysis overlapping with the IPF meta-analysis at Bonferroni threshold of P<0·0008.

| Variant | Gene | Ref/Alt | OR<sub>Alt</sub> (95% CI) |  |  | P |  |  |  |
| --- | --- | --- | --- | --- | --- | --- | --- | --- | --- |
|  |  |  | Finngen<br>Any cancer | UKB<br>Any cancer | Allen et al. (2020)<br>IPF | Finngen | UKB | Finngen &<br>UKB | Allen et al.<br>(2020) |
| rs116483731 | SPDL1 | G/A | 0.79 (0.76 – 0.83) | 0.89 (0.83 – 0.96) | 2.40 (1.70 – 3.40) | 1.08E-22 | 1.21E-03 | 2.10E-23 | 7.55E-07 |
| rs75691080 | RTEL1-STMN3 | C/T | 0.92 (0.9 – 0.94) | 0.94 (0.92 – 0.96) | 1.30 (1.16 – 1.46) | 8.68E-12 | 3.16E-07 | 5.23E-17 | 4.29E-06 |
| rs7902587 | OBFC1 | C/T | 1.05 (1.02 – 1.08) | 1.08 (1.06 – 1.11) | 0.76 (0.68 – 0.85) | 7.13E-04 | 1.54E-13 | 2.22E-15 | 2.08E-06 |
| rs2293607 | TERC-ACTRT3 | T/C | 0.95 (0.94 – 0.97) | 0.96 (0.95 – 0.98) | 1.29 (1.20 – 1.39) | 1.50E-07 | 1.95E-07 | 1.80E-13 | 2.33E-11 |
| rs10950456 | MAD1L1 | G/A | 0.97 (0.96 – 0.99) | 0.97 (0.96 – 0.98) | 1.20 (1.12 – 1.28) | 6.45E-04 | 4.41E-07 | 1.20E-09 | 1.19E-07 |
| rs112064988 | MAP2K1 | C/T | 1.03 (1.01 – 1.05) | 1.03 (1.02 – 1.05) | 0.87 (0.81 – 0.94) | 4.39E-04 | 1.26E-05 | 2.08E-08 | 6.77E-04 |

All loci have been previously described as cellular division related.^35–39^ *RTEL1-STMN3, TERC-ACTRT3* and *OBFC1* have been identified as both telomeres,^40–42^ and cancer associated. ^43–45^ Interestingly, *MAD1L1* was identified as IPF associated in the latest meta-analysis,^9^ however another variant was then reported as most likely causal.

### 3.8 SPDL1 is associated with cancer-related mosaic Chromosomal Alterations

A recent analysis on somatic loss of chromosome Y (LOY) in circulating white blood cells noted *SPDL1* as increasing the risk of LOY (rs116483731, odds ratio for the A allele 1·42; 95% CI 1·31 to 1·53; P=9·20×10^−18^) in the UKB.^46^ Hence, we first explored the frequency of mosaic Chromosomal Alterations (mCA, LOY being one of the most common type) in a subset of FinnGen population (n=148,272) with available intensity data from array genotyping, replicating the previously reported frequencies of mCA (Supplementary Table S8, Supplementary Fig. S4).^47^

We then assessed the variant-specific effects of the six cancer and IPF associated loci identified earlier (*SPDL1, MAD1L1, MAP2K1, RTEL1-STMN3, TERC-ACTRT3, OBFC1*) on accumulation of mCA. After adjusting for smoking status available for 49·5% of FinnGen participants, 73,339 participants were available for analysis.

Carrying the missense *SPDL1* risk allele for IPF (and protective of cancer) rs116483731 was seen protective (at P<6·9×10^−4^ after Bonferroni adjustment for multiple testing) for mCA event in any chromosome (odds ratio 0·71; 95% CI 0·63 to 0·79; P=5·36×10^−9^) and for both sex chromosomes, Table 4, beyond the protection of LOY shown in UKB.^46^

**Table 4.**
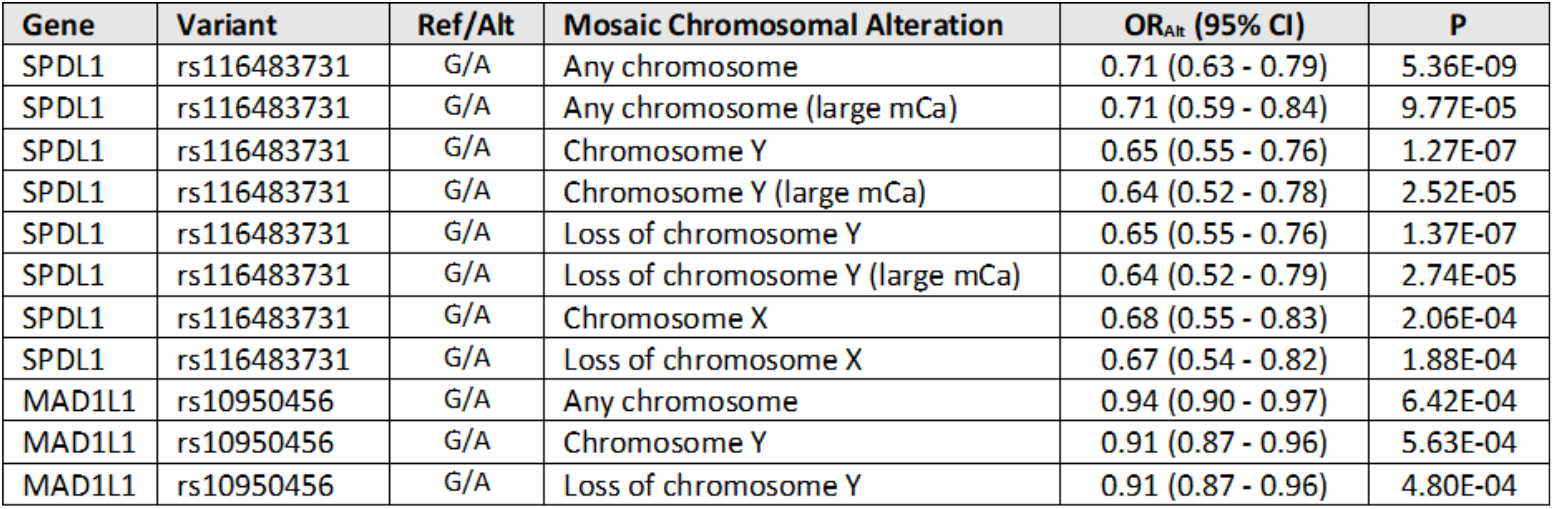
Variant-specific effects on mosaic chromosomal alteration in FinnGen reaching Bonferroni threshold of P<6·9×10^−4^.

Finally, we verified the effect of *MAD1L1* (rs10950456 allele A) in mCA accumulation (Table 4, Supplementary Table S8 with complete list of tested variants), highlighting that alternate alleles of both *MAD1L1* and *SPDL1* were found to decrease the risk of mCA in addition to lowering the risk of cancer.

## 4 DISCUSSION

The FinnGen study (in preparation) stands on two key strengths; i) the longitudinal data from multiple comprehensive nationwide health registries, and ii) the unique population history of concurrent bottlenecks and population growth, resulting in reduced allelic and locus heterogeneity causal to a disease.^48,49^ By utilizing health registers covering most aspects of human disease and health, we were able to assess extensively the beneficial or harmful effects a genetic variant might carry, considering even extremely rare alleles.

A missense variant in the *SPDL1* gene associated with increased risk of IPF was recently identified in FinnGen.^33^ Given that the variant is highly enriched in the Finnish population, it was not identified in the largest IPF-GWAS so far, although replicated. Strikingly in the present study, the variant was shown to protect against onset of cancer, while other coding alleles in the gene had no such effect. We further confirm the finding in a longitudinal analysis of cancer risk in FinnGen, also showing that protection from cancer is not due to increased risk of IPF.

We also identified two functional variants in the *TERT* gene independently associated with IPF. The first ever recurrent (non-familial, multiple occurrences in a population) LoF mutation and missense variant, both of which are polymorphic only in the Finnish, Swedish or Estonian populations. Both variants conferred an unusually high risk of IPF (accompanied by wide confidence intervals due to rarity of the alleles) and were also shown to protect against cancer onset, in addition to decreasing effects on the Lymphocyte Telomere Length.

We then performed a pan-cancer meta-analysis in FinnGen and UKB identifying 62 loci reaching genome-wide significance. Six lead variants were found to present opposite effects over IPF and cancer, while all loci have been associated with cellular division, three with telomere length.

Finally, consistent with the knowledge of chromosomal alterations – known to arise during cell division – we show that carrying a *SPDL1* or *MAD1L1* variant is associated with a markedly decreased number of mosaic chromosomal alterations, arising after fertilization. Notably, *MAD1L1* was earlier noted as IPF-associated in the latest meta-analysis.^9^ *SPDL1* has previously been described to act as a tumor suppressor in colorectal cancer,^50^ by controlling cell cycle progression.^51^

*MAD1L1* is one of the Mitotic Arrest Deficient (MAD) genes (and a homodimer of *MAD2L1*) constituting the Spindle Assembly Checkpoint (SAC) pathway ^52^ – in part regulated by *SPDL1* encoded protein.^37^ SAC can lead to mitotic arrest while maintaining genome stability and preventing chromosome mis-segregation and aneuploidy arising during cellular division. *MAD2L1* has been described as *BRCA1* associated in breast cancer and mesothelioma,^53,54^ while cancers related to *BRCA1/2* mutations are known to present more chromosomal instability. Furthermore, downregulation of *BRCA1* seems to inactivate SAC.^55,56^ SAC is activated and maintained (then leading to mitotic arrest and senescence or apoptosis) when mitotic spindle is compromised, in agreement with the opposite effects seen in IPF where the fibroblastic foci have been described even as completely lacking proliferating cells,^57^ suggesting that mitotic signaling and relevant regulation are key elements to both diseases.

The nature of the *SPDL1* missense variant is polarized in terms of human disease; only few examples of such antagonistic pleiotropy have been described until now. While it is associated with higher risk of rare IPF, it is protective of cancer – extremely common in any elderly population – all the while seeing no effect on other common diseases.

## Data Availability

FinnGen summary level results (summary statistics genome-wide association analysis and data at the phenotype level, including endpoint definitions, statistics about number of individuals, gender distribution, and longitudinal relationships) are made freely available every 6 month. The latest freely available data release IV consists of 176,899 participants.

https://finngen.gitbook.io/documentation/

## Contributors

JTK, PH, JMK, MK, and MJD were responsible for study-level analysis. AL, GG, MK, WZ, TP, JR, SS, TT, PP, JK, TL, MA, and AG were responsible for additional analysis and data management. JTK, AP, MM, and MJD were responsible design and funding. JTK, PH, AL, JP, GG, MA, MNMM, MK, WZ, JMK, TP, JR, SS, TT, PP, JK, MK, AG, AP, TL, MM, and MJD were responsible for critical review and writing of the manuscript.

