## Supplementary Appendix for "Genetic variant in *SPDL1* reveals novel mechanism linking pulmonary fibrosis risk and cancer protection"

**Disease endpoints in FinnGen registries**

IPF in FinnGen was defined by having an ICD-10 J84.1 diagnosis. Endpoints for specific neoplasias were constructed by compiling the data from cancer registry (ICD-O-3), hospital discharge registry (ICD-8 to 10 codes), and causes of death registry (ICD-8 to 10 codes). We limit the scope of this study to the four common cancers having at least 2000 cases in FinnGen. "Any cancer" endpoint includes all malignant neoplasms (ICD-8 to 10, and ICD-O-3).

**Array genotyping and imputation in FinnGen**

FinnGen participants were genotyped with Illumina and Affymetrix arrays (Illumina Inc., San Diego, and Thermo Fisher Scientific, Santa Clara, CA, USA), and genotype calls were made with the GenCall or zCall (for Illumina) and the AxiomGT1 algorithm for Affymetrix data.

Participants with ambiguous gender, high genotype missingness (>5%), excess heterozygosity (+-4SD) and non-Finnish ancestry were excluded; as well as all variants with high missingness (>2%), low HWE P-value (<1e-6) and minor allele count (MAC<3). Array data pre-phasing was carried out with Eagle 2.3.511 with the number of conditioning haplotypes set to 20,000.

Genotype imputation was done with Beagle 4.1,[1,2] (described in https://dx.doi.org/10.17504/protocols.io.xbgfijw) by using the SISu v3 population specific reference panel developed from high-quality data for 3,775 high-coverage (25-30x) WGS in Finns.

**Longitudinal analysis in FinnGen**

National registers cover the whole health history of study participants regardless of the age of study enrollment. To examine prevalence prior to joining the study, we followed participants from their birth till the age of study enrollment, IPF, or any cancer, whichever occurred first. To model incidence, we left-truncated the data at the age of study entry, i.e. followed participants from the age of study enrollment till IPF, any cancer, death, or end of the study follow-up, and excluded all participants with prevalent cancer or IPF prior to joining the study. In the combined analysis, we included all individuals at their birth and followed them to the end of the study follow-up or first event of interest, without considering the study recruitment time. We used age as the time-scale in all disease onset models. In contrast to case-control GWAS, participants with other cancer were not excluded but censored at the age of the first competing risk event to model cause-specific hazards.

Both genetic principal component covariates manifested non-linearity with any cancer onset (p<2e-16), and were thus included as penalized smoothing splines into all models. Proportional hazards assumption was evaluated using Schoenfeld residuals and respective proportional hazard tests in each model. After accounting for multiple testing, the first genomic principal component showed non-proportionality in respect to any cancer onset among prevalent cases (p=0.002). Consequently, we included both principal components as time-dependent covariates to all models for consistency.

Cumulative incidence functions were estimated with Aalen-Johansen estimators to account for competing risks of IPF, any cancer, and death without cancer. Illustrations of cumulative incidence are graphically smoothed to retain anonymity of study participants where appropriate.

All longitudinal statistical analyses were done using *R-4.0.3* and package *survival*.

**Calling somatic variants from array intensity data in FinnGen**

The genotype and intensity data was processed with the apt-probeset-genotype tool (http://www.affymetrix.com/support/developer/powertools/changelog/apt-probeset-genotype.html) and used to call mosaic chromosomal alterations (mCA) in 153,548 FinnGen (Release V) participants genotyped with FinnGen ThermoFisher Axiom custom array version 1 or 2. The calling was performed using the "txt" mode of the MoChA WDL pipeline (https://github.com/freeseek/mocha) that includes (i) converting intensity data to VCF files with B Allele Frequency (BAF) and Log R Ratio (LRR) using the gtc2vcf tool (https://github.com/freeseek/gtc2vcf), (ii) phasing the full cohort using the SHAPEIT4 software,[1] and (iii) calling mCA by detecting allelic imbalance of heterozygous sites based on LRR and phased BAF.

Participants with poor genotyping quality (185 participants with possible DNA contamination and 83 participants with discordance in phenotype-genotype sex) and event calls likely to be constitutional duplications or deletions were filtered using the criteria suggested by the MoChA pipeline. After further removing 1st and 2nd-degree relatives, a total of 148,272 participants were left for further analyses.

We analyzed mCa from autosomes and sex choromosomes separately.[3] Given MoChA was designed to search for allelic imbalance in heterozygous sites, only pseudoautosomal regions (PAR1 and PAR2) were analyzed for men by ignoring all non-PAR regions and then defining any mosaic events as mosaic Y rather than mosaic X. Thus, mosaic X is only defined for women and mosaic Y only for men. Given the most common mCa types in sex chromosomes are loss events for mLOY and mLOX,[4,5] we further defined two additional mCA subtypes specific for loss events based on the length of mCA events: (i) the mosaic loss of Y for mCA longer than 1Mb and (ii) the mosaic loss of X for mCA longer than 50 Mb.

**Telomere length analysis in the Northern Finland Birth Cohort and Finnish Twin Cohort**

The Northern Finland Birth Cohort 1966 (NFBC1966) is a prospective birth cohort study that recruited all pregnant women living in the Oulu and Lapland provinces of Finland with expected delivery in 1966, comprising in total 12,231 children and their parents with follow-ups at 1, 14, 31, and 46 years of age (University of Oulu 1966). All participants included in the study are from Finnish ancestry and are independent from the FinnGen sample. Mean relative leukocyte telomere length (LTL) was measured using monochrome multiplex qPCR method at 31-year follow-up. LTL at 46-year follow-up was measured in triplicates using albumin as a single-copy gene reference. The associations with genetic variants were assessed after a rank-based inverse normal transformation of LTL among 4,961 (48·4% males) and 3,547 (43·9% males) individuals at 31 and 46 years respectively.

Relative telomere length in the Finnish Twin Cohort (FTC) was measured with qPCR at the age of DNA collection among adult twins from three substudies: (i) the Nicotine Addiction Genetics study, (ii) the Finnish Twin Study on Ageing, and (iii) the TwinFat study.[6,7] The association of genetic variants and normalized logarithm of LTL was assessed using a linear mixed-model adjusted with age and sex, and by including family-identifier as a random effect to account for within-pair dependency.

#### Supplementary Results

#### IPF-GWAS in FinnGen

In addition to *SPDL1* and *TERT* variants, we identified two additional novel loci (at *TACC2* and *AXIN1*) at P<5×10$^{-8}$ in FinnGen. The identified *TACC2* locus at 10q26.13 included a single credible set where the most probable causal allele (rs967235139 causal probability 36·7%, MAF 0·21%, odds ratio for the C allele, 35·86; 95% CI 9·96 to 129·05; P=4·30×10$^{-8}$) is only polymorphic in the Finnish population according to the GnomAD resource,[8] and hence could not be replicated in the IPF meta-analysis with participants of non-Finnish European ancestry and considering only variants with MAF >1%.

Also the *AXIN1* locus at 16p13.3 included a single credible set for the locus (rs185488877 causal probability 32·5%, MAF 1·89%, odds ratio for the T allele, 2·74; 95% CI 1·93 to 3·91; P=2·32×10$^{-8}$) is rarely seen polymorphic in other than Finnish population as MAF in non-Finnish Europeans in gnomAD is 0·31% and the variant was not available in the IPF meta-analysis summary statistics. Axin1/2 have been previously described as cancer associated but also with Wnt-signaling,[9] relevant to fibrotic processes.

A Manhattan plot for test statistics from the FinnGen IPF is depicted in Supplementary Figure S1, in addition to quantile-quantile plot of observed and expected test statistics in Supplementary Figure S2. An overall genomic inflation factor ($\lambda_{GC}$) of 1·0311 was noted.

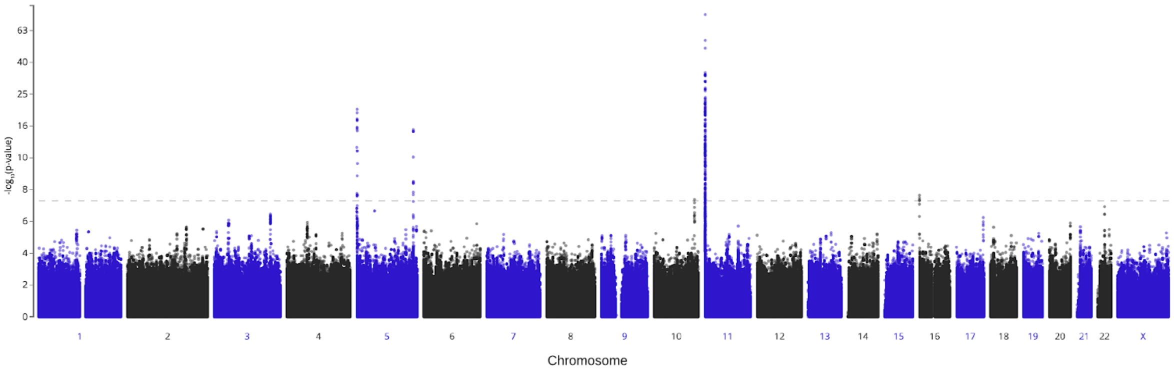


Supplementary Figure S1. Manhattan plot of the FinnGen IPF GWAS statistics. The dash line indicates the genome-wide significance threshold of a P value less than 5×10$^{-8}$.

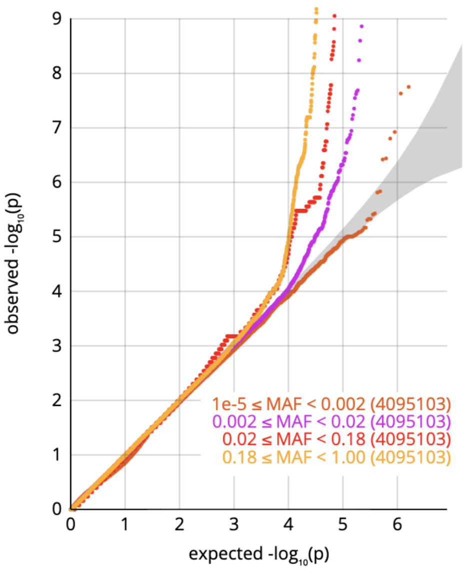


Supplementary Figure S2. Quantile-quantile plot of Observed and Expected test statistics in equally sized variant frequency bins in the FinnGen IPF GWAS.

#### Effects of SPDL1 missense variant in FinnGen

The potential pleiotropic effects of *SPDL1* missense variant (rs116483731) was assessed across FinnGen endpoints (n=2,925) and depicted in the table S1. Effects beyond $P<1\times10^{-5}$ were only seen in IPF, closely related pulmonary phenotypes. and different malignancy-related phenotypes.

| Endpoint | OR | P |
|---|---|---|
| Idiopathic pulmonary fibrosis | 3.13 | 9.97E-16 |
| Malignant neoplasm | 0.81 | 2.05E-15 |
| ILD, hospital admissions 1, main diag only | 2.25 | 4.48E-14 |
| Other respiratory diseases principally affecting the interstitium | 1.95 | 1.43E-13 |
| Interstitial lung disease | 2.09 | 2.10E-13 |
| Other insterstitial pulmonary diseases | 2.06 | 4.03E-13 |
| ILD, hospital admissions | 2.08 | 4.44E-13 |
| Respiratory diseases principally affecting the interstitium, IBD co-morbidites | 1.88 | 8.07E-11 |
| Malignant neoplasm of prostate (all cancers excluded) | 0.69 | 1.82E-08 |
| Malignant neoplasm of skin (all cancers excluded) | 0.77 | 2.10E-08 |
| Other malignant neoplasms of skin (=non-melanoma skin cancer) (all cancers excluded) | 0.77 | 2.23E-08 |
| malignant neoplasm of male genital organs (all cancers excluded) | 0.70 | 2.23E-08 |
| Malignant neoplasm of prostate | 0.72 | 2.37E-07 |
| ILD, hospital admission 2, with pulmonary infections | 1.91 | 2.83E-07 |
| malignant neoplasm of male genital organs | 0.73 | 3.26E-07 |
| Interstitial lung disease endpoints | 1.17 | 9.16E-07 |
| Malignant neoplasm of skin | 0.80 | 1.37E-06 |
| Other malignant neoplasms of skin (=non-melanoma skin cancer) | 0.80 | 1.41E-06 |
| ILD, hospital admissions 3, with pneumonia sepsis | 1.93 | 1.9E-06 |
| Malignant neoplasm of breast (all cancers excluded) | 0.79 | 7.15E-06 |

Supplementary Table S1. Disease and health related endpoint associations of *SPDL1* missense variant rs116483731 alternative allele A in FinnGen with $P<1\times10^{-5}$.

#### Effects of other SPDL1 functional variation in FinnGen

Altogether 13 predicted coding variants were seen in the *SPDL1* gene in FinnGen. Coding variation beyond rs116483731 was assessed within the scope of FinnGen endpoints, Supplementary Table S2. No other predicted functional variant associated with disease endpoints at $P<1x10^{-5}$.

| Variant | Ref/Alt | $AF_{alt}$ | INFO | Predicted effect |
|---|---|---|---|---|
| rs3797713 | T/C | 0.67 | 1.00 | Missense |
| rs3777084 | T/C | 0.67 | 1.00 | Missense |
| rs140884129 | G/A | 2.80E-03 | 0.99 | Missense |
| rs199675882 | A/G | 3.30E-03 | 1.00 | Missense |
| rs146344218 | C/T | 2.20E-03 | 0.96 | Missense |
| rs140005442 | A/G | 8.00E-04 | 0.99 | Splice acceptor |
| rs144297297 | G/A | 4.40E-04 | 0.95 | Missense |
| rs143453400 | A/T | 1.30E-03 | 0.95 | Missense |
| rs78480498 | C/A | 9.40E-03 | 0.99 | Missense |
| rs140080951 | G/A | 6.10E-03 | 0.99 | Missense |
| rs147788748 | C/T | 6.40E-04 | 0.97 | Missense |
| rs144151613 | A/T | 1.30E-03 | 0.97 | Missense |

Supplementary Table S2. Functional variation and associated disease endpoints ($P<1x10^{-4}$) in FinnGen.

##### *SPDL1* missense variant and the risk of IPF and cancer in FinnGen

To further assess the risk-increasing effect of rs116483731 in IPF and decreasing effect in cancer subtypes, we analyzed the FinnGen data with cause-specific Cox proportional hazards models separating participants by the disease status at study entry, referred as incident, prevalent and as a combination of both (Supplementary Table S3), and by considering other cancer and IPF as competing risk events.

| | Prevalent and incident | | Prevalent only | | Incident only | |
|---|---|---|---|---|---|---|
| **Disease** | **HR (95% CI)** | **p** | **HR (95% CI)** | **p** | **HR (95% CI)** | **p** |
| Colorectal cancer | 0.93 (0.79-1.11) | 0.44 | 0.87 (0.70-1.09) | 0.23 | 1.05 (0.80-1.38) | 0.74 |
| Prostate cancer | 0.71 (0.62-0.81) | 6.11e-07 | 0.74 (0.62-0.87) | 2.62e-04 | 0.67 (0.53-0.84) | 4.67e-04 |
| Breast cancer | 0.81 (0.73-0.90) | 5.55e-05 | 0.80 (0.71-0.90) | 3.01e-04 | 0.84 (0.69-1.01) | 0.07 |
| Nonmelanoma skin cancer | 0.82 (0.75-0.90) | 4.16e-05 | 0.79 (0.71-0.89) | 8.62e-05 | 0.88 (0.75-1.03) | 0.12 |
| Any cancer | 0.83 (0.79-0.87) | 4.23e-15 | 0.83 (0.79-0.88) | 1.52e-10 | 0.83 (0.76-0.90) | 5.59e-06 |
| IPF | 2.27 (1.86-2.76) | 4.46e-16 | 2.52 (1.98-3.22) | 1.08e-13 | 1.88 (1.35-2.64) | 2.22e-04 |

Supplementary Table S3. Hazard ratios in IPF, cancer, and subtypes from cause-specific Cox proportional hazards models for the *SPDL1* missense variant rs116483731 allele A in FinnGen.

##### Association of *SPDL1* and *TERT* variants with telomere length in NFBC1966 and FTC

The association of three identified functional variants in *SPDL1* and *TERT* on Lymphocyte Telomere Length (LTL) was analyzed in the independent NFBC1966 study at the age of 31 and 46. We saw no effect for the *SPDL1* missense variant rs116483731, while association was observed for both of the *TERT* alleles rs770066110 and rs776981958 in NFBC1966 (Supplementary Table S4). The effect of both *TERT* variants was confirmed in the FTC study (Supplementary Table S5). A large LTL meta-analysis [10] further supports our notion that the SPDL1 missense variant seems to have no effect on the LTL (rs116483731 allele A, **β**=0·0099, P=0·21).

| Variant | Ref/Alt | LTL Age | $Beta_{alt}$ (95% CI) | P |
|---|---|---|---|---|
| rs770066110 | G/A | 31 | -0.66 (-0.57 – -0.10) | 0.021 |
| rs776981958 | T/C | 31 | -0.90 (-0.49 – -0.41) | 3.39E-04 |
| rs116483731 | G/A | 31 | 0.03 (-0.11 – 0.14) | 0.62 |
| rs770066110 | G/A | 46 | -0.79 (-0.65 – -0.14) | 0.017 |
| rs776981958 | T/C | 46 | -1.00 (-0.62 – -0.38) | 1.59E-03 |
| rs116483731 | G/A | 46 | 0.02 (-0.13 – 0.14) | 0.81 |

Supplementary **Table S4. Effect of *SPDL1* and *TERT* functional variants Lymphocyte Telomere Length (LTL) in the independent NFBC1966 study, measured at the age of 31yrs and 46yrs.** Linear regression of carrier status and normalized LTL (rank-based inverse normal transformation).

| Variant | Ref/Alt | $Beta_{alt}$ (95% CI) | P |
|---|---|---|---|
| rs770066110 | G/A | -2.41 (-3.86 – -0.96) | 1.17E-03 |
| rs776981958 | T/C | -0.90 (-1.66 – -0.15) | 0.019 |

**Supplementary Table** S5. **Effect of *SPDL1* and *TERT* functional variants Lymphocyte Telomere Length (LTL) in FTC study, measured at the time of DNA collection in each substudy.** Linear mixed-model regression of carrier status and normalized logarithm of LTL, models were adjusted with age and sex where within-pair dependency was accounted for by using family-identifier as a random effect.

#### *TERT* variants and the risk of IPF and cancer in FinnGen

We confirmed the increasing effect for IPF-risk and the decreasing effect for any cancer of the two functional *TERT* mutations (Supplementary Table S6). In addition, a protective effect against prostate cancer was observed.

| | Prevalent and incident | | Prevalent only | | Incident only | |
|---|---|---|---|---|---|---|
| **Disease** | **HR (95% CI)** | **p** | **HR (95% CI)** | **p** | **HR (95% CI)** | **p** |
| Colorectal cancer | 0.88 (0.44-1.76) | 0.72 | - | - | - | - |
| Prostate cancer | 0.28 (0.12-0.67) | 4.25e-3 | - | - | - | - |
| Breast cancer | 0.99 (0.69-1.40) | 0.94 | 0.98 (0.64-1.50) | 0.92 | 1.02 (0.55-1.9) | 0.96 |
| Nonmelanoma skin cancer | 0.80 (0.56-1.17) | 0.25 | 0.77 (0.49-1.23) | 0.28 | 0.86 (0.46-1.61) | 0.64 |
| Any cancer | 0.78 (0.65-0.94) | 8.36e-3 | 0.76 (0.60-0.95) | 0.02 | 0.84 (0.62-1.14) | 0.25 |
| IPF | 13.9 (10.3-18.8) | 3.00e-65 | 14.1 (9.59-20.7) | 1.34e-41 | 13.8 (8.41-22.5) | 1.68e-25 |

Supplementary Table S6. Hazard ratios of IPF, cancer, and subtypes from cause-specific Cox proportional hazards models in carriers of either of *TERT* functional mutations. Instances with less than five mutation-carrying cases were left out from the analysis.

#### Mosaic Chromosomal Alterations in FinnGen

Most of the mCA events were seen in the sex chromosomes and autosomal events accounted for 25·6% of all mCAs in women and 13·5% in men (Supplementary Table S7, Supplementary Fig. S3), consistent with previous reports.[11] The mosaic deletion in chromosome Y, usually termed as mosaic loss of chromosome Y (mLOY), was the most common form of clonal mosaicism observed. Among the 10,309 mCA events observed in chromosome Y with distinguishable types, 91·8% were classified as mLOY (Supplementary Table S7). Odds ratios of six variants associated with both IPF and cancer with their effects on the incidence of mCA events in FinnGen are illustrated in the Supplementary Table S8.

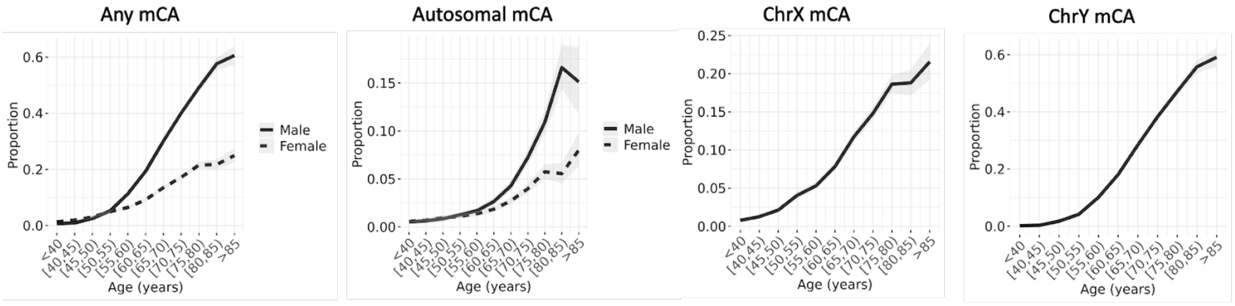


Supplementary Figure S3. Prevalence of mosaic chromosomal alteration (mCA) by 5-year age bins in male and female FinnGen participants (a subset with intensity data available, n=148,272).

| Mosaic Chromosomal Alteration | Women | Men | Total (%) |
|---|---|---|---|
| Any chromosome | 6,519 | 11,498 | **18,017 (48 %)** |
| Autosomal | 1,430 | 1,516 | **2,946 (7.9 %)** |
| Chromosome X | 5,309 | 0 | **5,309 (14 %)** |
| Chromosome Y | 0 | 10,449 | **10,449 (28 %)** |
| Autosomal and ChrX | 220 | 0 | **220 (0.59 %)** |
| Autosomal and ChrY | 0 | 467 | **467 (1.2 %)** |
| **Total (%)** | **13,478 (36 %)** | **23,930 (64 %)** | **37,408 (100 %)** |

Supplementary Table S7. Number mCA carriers in FinnGen by sex (in a subset with intensity data available, n=148,272).

| Mosaic Chromosomal Alteration | Rsid | Gene | Ref/Alt | $OR_{Alt}$ (95% CI) | P |
|---|---|---|---|---|---|
| AnyChr | rs2293607 | TERC-ACTRT3 | T/C | 1.00 (0.96 - 1.04) | 0.88 |
| AnyChr | rs116483731 | SPDL1 | G/A | 0.71 (0.63 - 0.79) | 5.36E-09 |
| AnyChr | rs10950456 | MAD1L1 | G/A | 0.94 (0.90 - 0.97) | 6.42E-04 |
| AnyChr | rs7902587 | OBFC1 | C/T | 0.96 (0.90 - 1.02) | 0.22 |
| AnyChr | rs112064988 | MAP2K1 | C/T | 1.03 (0.99 - 1.08) | 0.12 |
| AnyChr | rs75691080 | RTEL1-STMN3 | C/T | 0.97 (0.91 - 1.02) | 0.21 |
| Autosomes | rs2293607 | TERC-ACTRT3 | T/C | 0.98 (0.90 - 1.08) | 0.71 |
| Autosomes | rs116483731 | SPDL1 | G/A | 0.88 (0.68 - 1.11) | 0.3 |
| Autosomes | rs10950456 | MAD1L1 | G/A | 0.90 (0.82 - 0.97) | 8.10E-03 |
| Autosomes | rs7902587 | OBFC1 | C/T | 1.05 (0.91 - 1.21) | 0.46 |
| Autosomes | rs112064988 | MAP2K1 | C/T | 0.99 (0.90 - 1.09) | 0.83 |
| Autosomes | rs75691080 | RTEL1-STMN3 | C/T | 0.94 (0.83 - 1.07) | 0.34 |
| Large_AnyChr | rs2293607 | TERC-ACTRT3 | T/C | 1.02 (0.96 - 1.09) | 0.47 |
| Large_AnyChr | rs116483731 | SPDL1 | G/A | 0.71 (0.59 - 0.84) | 9.77E-05 |
| Large_AnyChr | rs10950456 | MAD1L1 | G/A | 0.94 (0.88 - 0.99) | 1.74E-02 |
| Large_AnyChr | rs7902587 | OBFC1 | C/T | 0.92 (0.83 - 1.01) | 0.08 |
| Large_AnyChr | rs112064988 | MAP2K1 | C/T | 1.06 (0.99 - 1.12) | 0.08 |
| Large_AnyChr | rs75691080 | RTEL1-STMN3 | C/T | 0.97 (0.89 - 1.05) | 0.47 |
| Large_Autosomes | rs2293607 | TERC-ACTRT3 | T/C | 0.97 (0.86 - 1.10) | 0.63 |
| Large_Autosomes | rs116483731 | SPDL1 | G/A | 0.98 (0.70 - 1.34) | 0.92 |
| Large_Autosomes | rs10950456 | MAD1L1 | G/A | 0.93 (0.83 - 1.05) | 0.23 |
| Large_Autosomes | rs7902587 | OBFC1 | C/T | 0.99 (0.81 - 1.20) | 0.94 |
| Large_Autosomes | rs112064988 | MAP2K1 | C/T | 1.00 (0.88 - 1.13) | 0.99 |
| Large_Autosomes | rs75691080 | RTEL1-STMN3 | C/T | 0.79 (0.65 - 0.94) | 1.15E-02 |
| ChrY | rs2293607 | TERC-ACTRT3 | T/C | 1.02 (0.96 - 1.07) | 0.6 |
| ChrY | rs116483731 | SPDL1 | G/A | 0.65 (0.55 - 0.76) | 1.27E-07 |
| ChrY | rs10950456 | MAD1L1 | G/A | 0.91 (0.87 - 0.96) | 5.63E-04 |
| ChrY | rs7902587 | OBFC1 | C/T | 0.92 (0.84 - 1.01) | 0.07 |
| ChrY | rs112064988 | MAP2K1 | C/T | 1.07 (1.01 - 1.13) | 2.56E-02 |
| ChrY | rs75691080 | RTEL1-STMN3 | C/T | 0.97 (0.90 - 1.05) | 0.49 |
| Large_ChrY | rs2293607 | TERC-ACTRT3 | T/C | 1.03 (0.96 - 1.10) | 0.41 |
| Large_ChrY | rs116483731 | SPDL1 | G/A | 0.64 (0.52 - 0.78) | 2.52E-05 |
| Large_ChrY | rs10950456 | MAD1L1 | G/A | 0.93 (0.87 - 0.99) | 2.21E-02 |
| Large_ChrY | rs7902587 | OBFC1 | C/T | 0.88 (0.79 - 0.99) | 2.99E-02 |
| Large_ChrY | rs112064988 | MAP2K1 | C/T | 1.10 (1.03 - 1.18) | 7.50E-03 |
| Large_ChrY | rs75691080 | RTEL1-STMN3 | C/T | 1.02 (0.93 - 1.12) | 0.67 |
| LOY | rs2293607 | TERC-ACTRT3 | T/C | 1.02 (0.96 - 1.07) | 0.58 |
| LOY | rs116483731 | SPDL1 | G/A | 0.65 (0.55 - 0.76) | 1.37E-07 |
| LOY | rs10950456 | MAD1L1 | G/A | 0.91 (0.87 - 0.96) | 4.80E-04 |
| LOY | rs7902587 | OBFC1 | C/T | 0.92 (0.84 - 1.01) | 0.07 |
| LOY | rs112064988 | MAP2K1 | C/T | 1.07 (1.01 - 1.13) | 2.56E-02 |
| LOY | rs75691080 | RTEL1-STMN3 | C/T | 0.97 (0.90 - 1.05) | 0.49 |
| Large_LOY | rs2293607 | TERC-ACTRT3 | T/C | 1.03 (0.96 - 1.11) | 0.38 |
| Large_LOY | rs116483731 | SPDL1 | G/A | 0.64 (0.52 - 0.79) | 2.74E-05 |
| Large_LOY | rs10950456 | MAD1L1 | G/A | 0.93 (0.87 - 0.99) | 1.88E-02 |
| Large_LOY | rs7902587 | OBFC1 | C/T | 0.88 (0.79 - 0.99) | 3.29E-02 |
| Large_LOY | rs112064988 | MAP2K1 | C/T | 1.10 (1.03 - 1.18) | 7.35E-03 |
| Large_LOY | rs75691080 | RTEL1-STMN3 | C/T | 1.02 (0.93 - 1.12) | 0.67 |
| ChrX | rs2293607 | TERC-ACTRT3 | T/C | 0.99 (0.93 - 1.06) | 0.83 |
| ChrX | rs116483731 | SPDL1 | G/A | 0.68 (0.55 - 0.83) | 2.06E-04 |
| ChrX | rs10950456 | MAD1L1 | G/A | 0.98 (0.92 - 1.04) | 0.44 |
| ChrX | rs7902587 | OBFC1 | C/T | 0.97 (0.86 - 1.08) | 0.52 |
| ChrX | rs112064988 | MAP2K1 | C/T | 1.02 (0.95 - 1.09) | 0.56 |
| ChrX | rs75691080 | RTEL1-STMN3 | C/T | 0.96 (0.87 - 1.05) | 0.38 |
| Large_ChrX | rs2293607 | TERC-ACTRT3 | T/C | 1.08 (0.85 - 1.36) | 0.55 |
| Large_ChrX | rs116483731 | SPDL1 | G/A | 0.55 (0.22 - 1.12) | 0.14 |
| Large_ChrX | rs10950456 | MAD1L1 | G/A | 0.93 (0.75 - 1.16) | 0.49 |
| Large_ChrX | rs7902587 | OBFC1 | C/T | 1.01 (0.67 - 1.45) | 0.97 |
| Large_ChrX | rs112064988 | MAP2K1 | C/T | 0.76 (0.58 - 0.99) | 4.34E-02 |
| Large_ChrX | rs75691080 | RTEL1-STMN3 | C/T | 0.87 (0.60 - 1.22) | 0.43 |
| LOX | rs2293607 | TERC-ACTRT3 | T/C | 1.00 (0.93 - 1.07) | 0.88 |
| LOX | rs116483731 | SPDL1 | G/A | 0.67 (0.54 - 0.82) | 1.88E-04 |
| LOX | rs10950456 | MAD1L1 | G/A | 0.97 (0.91 - 1.04) | 0.42 |
| LOX | rs7902587 | OBFC1 | C/T | 0.96 (0.86 - 1.07) | 0.47 |
| LOX | rs112064988 | MAP2K1 | C/T | 1.03 (0.96 - 1.11) | 0.39 |
| LOX | rs75691080 | RTEL1-STMN3 | C/T | 0.96 (0.87 - 1.06) | 0.42 |
| Large_LOX | rs2293607 | TERC-ACTRT3 | T/C | 1.21 (0.89 - 1.62) | 0.22 |
| Large_LOX | rs116483731 | SPDL1 | G/A | 0.15 (0.01 - 0.67) | 0.06 |
| Large_LOX | rs10950456 | MAD1L1 | G/A | 0.98 (0.74 - 1.31) | 0.88 |
| Large_LOX | rs7902587 | OBFC1 | C/T | 0.95 (0.56 - 1.52) | 0.84 |
| Large_LOX | rs112064988 | MAP2K1 | C/T | 0.78 (0.55 - 1.08) | 0.14 |
| Large_LOX | rs75691080 | RTEL1-STMN3 | C/T | 0.93 (0.58 - 1.41) | 0.75 |

Supplementary Table S8. Odds ratios of 6 variants found to be associated with both IPF and cancer, with their effects on the incidence of different mCA events in FinnGen participants. A logistic regression adjusted with age, $age^2$, sex and ever-smoking status (in a subset with intensity data and smoking status available, n=73.339)

### SPDL1 expression in TCGA

We compared the expression of *SPDL1* between normal and tumor samples in the TCGA data [12]. The results shown here are in whole or in part based upon data generated by the TCGA Research Network (https://www.cancer.gov/tcga). Altogether 11 cancer types having RNA-sequencing data from at least 20 normal tissue samples were included in the analysis: Breast invasive carcinoma (BRCA), Head and Neck squamous cell carcinoma (HNSC), Kidney Chromophobe (KICH), Kidney renal clear cell carcinoma (KIRC), Kidney renal papillary cell carcinoma (KIRP), Liver hepatocellular carcinoma (LIHC), Lung adenocarcinoma (LUAD), Lung squamous cell carcinoma (LUSC), Prostate adenocarcinoma (PRAD), Stomach adenocarcinoma (STAD), and Thyroid carcinoma (THCA) (Supplementary Table S9).

| Cancer Type | Normal Sample Count | Tumor Sample Count | P |
|---|---|---|---|
| Breast invasive carcinoma (BRCA) | 112 | 112 | 3.9e-13 |
| Head and Neck squamous cell carcinoma (HNSC) | 43 | 43 | 7.71e-03 |
| Kidney Chromophobe (KICH) | 25 | 25 | 1.28e-01 |
| Kidney renal clear cell carcinoma (KIRC) | 72 | 72 | 1.97e-07 |
| Kidney renal papillary cell carcinoma (KIRP) | 32 | 32 | 6.66e-04 |
| Liver hepatocellular carcinoma (LIHC) | 50 | 50 | 4.13e-08 |
| Lung adenocarcinoma (LUAD) | 58 | 58 | 8.69e-10 |
| Lung squamous cell carcinoma (LUSC) | 51 | 51 | 3.58e-10 |
| Prostate adenocarcinoma (PRAD) | 52 | 52 | 2.4e-03 |
| Stomach adenocarcinoma (STAD) | 32 | 32 | 1.49e-06 |
| Thyroid carcinoma (THCA) | 59 | 59 | 2.49e-01 |

Supplementary Table S9. Comparison of Normalized *SPDL1* expression in TCGA tumors and normal samples.

Cancer cohorts were subsampled to keep the same individuals in normal and tumor groups and a t-test was used to compare normalized gene expression levels between the two tissue source sites for each cancer type (Supplementary Fig. S4). Out of tested cancer types, 9 demonstrated significantly higher expression of *SPDL1* in tumors rather than in normal tissue samples (at threshold $p<4{\cdot}5x10^{-3}$).

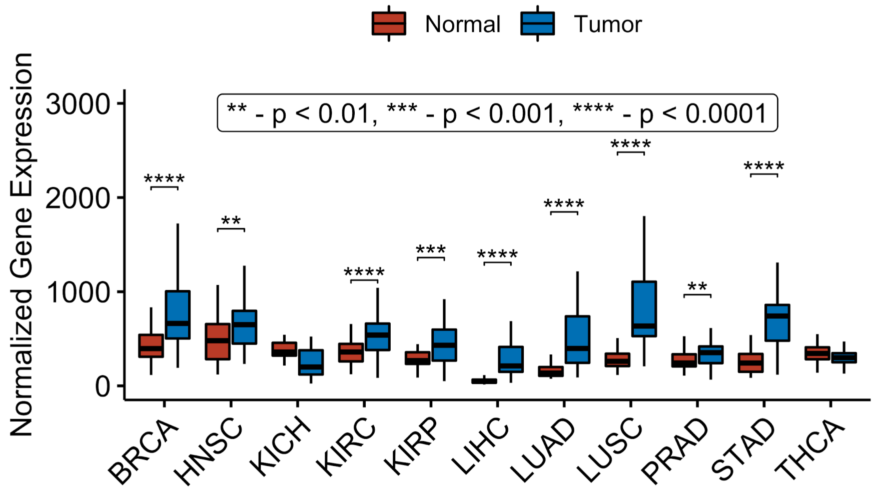


Supplementary Figure S4. Normalized *SPDL1* expression in TCGA tumors and normal samples.

#### Overlap of IPF meta-analysis with FinnGen and UKB pan-cancer analysis

Leading 62 SNVs from the FinnGen and UKB pan-cancer analysis and their overlap with the latest IPF meta-analysis (Allen et al. 2020) is illustrated in the Supplementary Table S10.

| | | | $OR_{Alt}$ (95% CI) | | | P | | | |
|---|---|---|---|---|---|---|---|---|---|
| **Variant** | **Gene** | **Ref/Alt** | **Finngen Any cancer** | **UKBB Any cancer** | **Allen et al. (2020) IPF** | **Finngen** | **UKBB** | **Finngen & UKBB** | **Allen et al. (2020)** |
| rs2293607 | ACTRT3 | T/C | 0.95 (0.94 - 0.97) | 0.96 (0.95 - 0.98) | 1.29 (1.20 - 1.39) | 1.50E-07 | 1.95E-07 | 1.80E-13 | 2.33E-11 |
| rs10950456 | MAD1L1 | G/A | 0.97 (0.96 - 0.99) | 0.97 (0.96 - 0.98) | 1.20 (1.12 - 1.28) | 6.45E-04 | 4.41E-07 | 1.20E-09 | 1.19E-07 |
| rs116483731 | SPDL1 | G/A | 0.79 (0.76 - 0.83) | 0.89 (0.83 - 0.96) | 2.40 (1.70 - 3.40) | 1.08E-22 | 0.00121 | 2.10E-23 | 7.55E-07 |
| rs7902587 | OBFC1 | C/T | 1.05 (1.02 - 1.08) | 1.08 (1.06 - 1.11) | 0.76 (0.68 - 0.85) | 7.13E-04 | 1.54E-13 | 2.22E-15 | 2.08E-06 |
| rs75691080 | STMN3 | C/T | 0.92 (0.9 - 0.94) | 0.94 (0.92 - 0.96) | 1.30 (1.16 - 1.46) | 8.68E-12 | 3.16E-07 | 5.23E-17 | 4.29E-06 |
| rs112064988 | MAP2K1 | C/T | 1.03 (1.01 - 1.05) | 1.03 (1.02 - 1.05) | 0.87 (0.81 - 0.94) | 4.39E-04 | 1.26E-05 | 2.08E-08 | 6.77E-04 |
| rs17599629 | CERS2 | A/G | 1.05 (1.03 - 1.07) | 1.04 (1.03 - 1.06) | 1.14 (1.05 - 1.23) | 4.55E-06 | 1.53E-08 | 3.35E-13 | 1.32E-03 |
| rs3818625 | SMC2 | G/A | 1.04 (1.02 - 1.06) | 1.03 (1.02 - 1.04) | 1.09 (1.02 - 1.16) | 6.92E-07 | 4.51E-06 | 2.43E-11 | 0.01 |
| rs12203592 | IRF4 | C/T | 1.12 (1.07 - 1.17) | 1.13 (1.11 - 1.15) | 1.09 (1.00 - 1.19) | 1.01E-06 | 3.71E-55 | 2.51E-60 | 0.04 |
| rs78378222 | TP53 | T/G | 1.24 (1.17 - 1.32) | 1.25 (1.18 - 1.33) | 0.71 (0.52 - 0.98) | 1.40E-12 | 9.84E-15 | 9.67E-26 | 0.04 |
| rs801108 | RP4-799G3.2 | C/G | 1.04 (1.02 - 1.06) | 1.07 (1.05 - 1.08) | 1.08 (1.00 - 1.17) | 9.90E-06 | 1.22E-18 | 7.76E-22 | 0.04 |
| rs11611584 | KRT5 | C/A | 1.04 (1.02 - 1.07) | 1.06 (1.04 - 1.08) | 0.89 (0.80 – 1.00) | 6.47E-04 | 1.07E-07 | 5.10E-10 | 0.05 |
| rs37004 | N/A | C/T | 0.96 (0.95 - 0.98) | 0.96 (0.94 - 0.97) | 1.07 (0.99 - 1.16) | 6.61E-05 | 1.76E-09 | 6.12E-13 | 0.08 |
| rs2384054 | N/A | T/C | 0.97 (0.95 - 0.98) | 0.98 (0.96 - 0.99) | 1.06 (0.99 - 1.13) | 2.54E-05 | 0.000249 | 4.01E-08 | 0.09 |
| rs6059655 | RALY | A/G | 0.95 (0.90 – 1.00) | 0.89 (0.87 - 0.91) | 0.91 (0.81 - 1.02) | 3.68E-02 | 3.33E-28 | 3.98E-28 | 0.10 |
| rs148297846 | TERT | G/A | 1.06 (1.02 - 1.11) | 1.07 (1.05 - 1.09) | 0.91 (0.80 - 1.03) | 7.05E-03 | 6.17E-09 | 1.51E-10 | 0.12 |
| rs17879961 | CHEK2 | A/G | 1.17 (1.12 - 1.23) | 1.49 (1.08 - 2.06) | 0.56 (0.26 - 1.24) | 2.99E-12 | 0.0155 | 4.40E-13 | 0.15 |
| rs7028268 | CDKN2B-AS1 | G/A | 0.96 (0.94 - 0.97) | 0.96 (0.94 - 0.97) | 0.95 (0.89 - 1.02) | 1.92E-08 | 1.93E-11 | 2.09E-18 | 0.16 |
| rs28792460 | RGS22 | A/G | 0.94 (0.92 - 0.96) | 0.95 (0.94 - 0.97) | 1.06 (0.97 - 1.16) | 1.85E-11 | 4.46E-08 | 9.08E-18 | 0.19 |
| rs340797 | N/A | G/T | 0.97 (0.94 – 1.00) | 0.95 (0.93 - 0.96) | 0.93 (0.84 - 1.04) | 3.02E-02 | 5.54E-08 | 1.04E-08 | 0.21 |
| rs72928038 | BACH2 | G/A | 0.96 (0.94 - 0.98) | 0.95 (0.94 - 0.97) | 0.95 (0.87 - 1.03) | 5.61E-04 | 1.04E-08 | 2.44E-11 | 0.24 |
| rs8037137 | PRC1 | T/C | 0.94 (0.92 - 0.96) | 0.96 (0.94 - 0.98) | 1.05 (0.96 - 1.16) | 2.81E-09 | 3.82E-06 | 1.52E-13 | 0.28 |
| rs13316357 | FOXP1 | A/G | 1.04 (1.02 - 1.06) | 1.04 (1.02 - 1.05) | 0.96 (0.90 - 1.03) | 6.08E-06 | 1.20E-07 | 3.41E-12 | 0.29 |
| rs157936 | KLF14, MIR29A | T/G | 0.97 (0.96 - 0.99) | 0.96 (0.95 - 0.98) | 0.96 (0.90 - 1.03) | 2.05E-04 | 1.41E-07 | 1.45E-10 | 0.30 |
| rs55676236 | NEK10 | A/C | 0.97 (0.95 - 0.98) | 0.97 (0.96 - 0.98) | 1.03 (0.97 - 1.11) | 3.22E-05 | 6.58E-07 | 9.12E-11 | 0.32 |
| rs1045020 | SLC22A5 | C/T | 0.97 (0.96 - 0.99) | 0.95 (0.93 - 0.97) | 1.05 (0.95 - 1.16) | 4.94E-03 | 1.59E-08 | 2.35E-09 | 0.33 |
| rs62202836 | N/A | C/T | 0.96 (0.94 - 0.98) | 0.97 (0.96 - 0.98) | 1.04 (0.96 - 1.12) | 4.35E-06 | 6.52E-05 | 1.82E-09 | 0.33 |
| rs661204 | N/A | G/A | 1.06 (1.03 - 1.08) | 1.06 (1.04 - 1.08) | 0.95 (0.86 - 1.05) | 1.02E-06 | 2.56E-10 | 1.50E-15 | 0.33 |
| rs72755295 | EXO1 | A/G | 1.17 (1.12 - 1.22) | 1.07 (1.04 - 1.11) | 0.91 (0.74 - 1.13) | 2.32E-12 | 5.21E-05 | 5.14E-14 | 0.39 |
| rs149934734 | ATM | C/T | 1.27 (1.17 - 1.38) | 1.09 (1.05 - 1.13) | 0.91 (0.74 - 1.13) | 2.01E-08 | 2.74E-05 | 5.90E-10 | 0.40 |
| rs35138525 | N/A | C/T | 1.04 (1.02 - 1.05) | 1.03 (1.01 - 1.04) | 1.03 (0.96 - 1.10) | 2.48E-05 | 5.31E-05 | 7.78E-09 | 0.41 |
| rs11599804 | FGFR2 | G/A | 1.05 (1.04 - 1.07) | 1.05 (1.03 - 1.06) | 1.03 (0.96 - 1.10) | 2.14E-10 | 6.12E-12 | 9.87E-21 | 0.43 |
| rs4784227 | CASC16 | C/T | 1.05 (1.03 - 1.06) | 1.05 (1.04 - 1.07) | 0.97 (0.90 - 1.05) | 9.19E-07 | 6.50E-13 | 4.31E-18 | 0.45 |
| rs62237617 | CHEK2 | C/T | 1.74 (1.59 - 1.90) | 1.67 (1.46 - 1.90) | 0.73 (0.33 - 1.64) | 1.06E-33 | 2.95E-14 | 3.00E-46 | 0.45 |
| rs35251485 | ZFHX4-AS1 | G/A | 0.92 (0.89 - 0.95) | 0.90 (0.88 - 0.92) | 0.96 (0.84 - 1.10) | 3.00E-08 | 4.38E-15 | 1.26E-21 | 0.53 |
| rs11263761 | HNF1B | A/G | 0.96 (0.94 - 0.97) | 0.97 (0.96 - 0.98) | 0.98 (0.92 - 1.05) | 2.62E-07 | 1.02E-05 | 3.01E-11 | 0.57 |
| rs162892 | P4HA2 | A/G | 1.03 (1.01 - 1.04) | 1.03 (1.02 - 1.05) | 0.98 (0.91 - 1.05) | 8.15E-04 | 4.92E-06 | 1.65E-08 | 0.58 |
| rs188140481 | N/A | T/A | 1.36 (1.27 - 1.45) | 1.29 (1.19 - 1.40) | 0.86 (0.50 - 1.48) | 3.61E-20 | 1.69E-09 | 6.18E-28 | 0.59 |
| rs112381112 | TANGO6 | G/T | 0.95 (0.93 - 0.98) | 0.94 (0.92 - 0.97) | 1.03 (0.9 - 1.18) | 7.92E-04 | 8.23E-06 | 2.89E-08 | 0.67 |
| rs12134662 | PADI6 | A/G | 1.04 (1.02 - 1.06) | 1.06 (1.05 - 1.08) | 1.01 (0.95 - 1.08) | 1.48E-06 | 1.41E-21 | 1.64E-25 | 0.70 |
| rs11571815 | BRCA2 | G/A | 1.18 (1.09 - 1.28) | 1.18 (1.10 - 1.26) | 1.07 (0.74 - 1.56) | 4.57E-05 | 7.41E-07 | 1.43E-10 | 0.72 |
| rs12653202 | N/A | A/C | 1.04 (1.02 - 1.07) | 1.04 (1.02 - 1.06) | 1.01 (0.92 - 1.11) | 2.38E-04 | 1.02E-05 | 9.73E-09 | 0.83 |
| rs55775505 | CASC15 | C/T | 1.04 (1.02 - 1.06) | 1.03 (1.02 - 1.05) | 0.99 (0.93 - 1.07) | 3.21E-06 | 1.44E-06 | 2.87E-11 | 0.85 |
| rs113934718 | ATAD5 | C/A | 0.98 (0.96 - 0.99) | 0.97 (0.95 - 0.98) | 0.99 (0.92 - 1.07) | 4.35E-03 | 1.11E-06 | 2.47E-08 | 0.87 |
| rs4268748 | DEF8 | T/C | 1.06 (1.05 - 1.08) | 1.07 (1.06 - 1.09) | 1.00 (0.93 - 1.07) | 9.56E-12 | 2.48E-24 | 2.14E-34 | 0.89 |
| rs4848599 | N/A | T/C | 1.06 (1.03 - 1.09) | 1.05 (1.03 - 1.07) | 0.99 (0.90 - 1.10) | 9.49E-05 | 3.50E-06 | 1.61E-09 | 0.91 |
| rs1126809 | TYR | G/A | 1.06 (1.03 - 1.08) | 1.06 (1.04 - 1.07) | 1.00 (0.93 - 1.08) | 1.12E-07 | 2.96E-16 | 1.95E-22 | 0.93 |
| rs214793 | TGM3 | C/T | 0.95 (0.93 - 0.97) | 0.93 (0.92 - 0.95) | 1.00 (0.92 - 1.09) | 4.95E-06 | 2.00E-17 | 1.40E-21 | 0.94 |
| rs719338 | ZMIZ1 | G/T | 0.97 (0.96 - 0.99) | 0.97 (0.96 - 0.98) | 1.00 (0.94 - 1.07) | 1.27E-03 | 2.66E-06 | 1.31E-08 | 0.95 |
| rs62329727 | N/A | T/C | 1.12 (0.97 - 1.30) | 1.19 (1.12 - 1.26) | 1.00 (0.72 - 1.40) | 1.16E-01 | 4.96E-09 | 1.85E-09 | 0.99 |
| rs79134926 | N/A | G/T | 0.97 (0.95 - 0.99) | 0.92 (0.90 - 0.94) | 1.00 (0.90 - 1.11) | 1.56E-03 | 2.66E-16 | 3.61E-16 | 1.00 |
| rs138213197 | HOXB13 | C/T | 1.53 (1.40 - 1.67) | 1.49 (1.28 - 1.73) | N/A | 6.43E-21 | 1.54E-07 | 6.14E-27 | N/A |
| rs148874756, rs201616617 | N/A | C/CTGT | 0.97 (0.95 - 0.99) | 0.97 (0.96 - 0.98) | N/A | 3.63E-04 | 1.79E-05 | 2.47E-08 | N/A |
| rs34138847, rs548053708 | N/A | GTATT/G | 1.04 (1.02 - 1.05) | 1.03 (1.01 - 1.04) | N/A | 2.53E-05 | 5.84E-05 | 7.97E-09 | N/A |
| rs515726080 | PALB2 | A/G | 3.56 (2.84 - 4.47) | N/A | N/A | 6.76E-28 | N/A | 6.76E-28 | N/A |
| rs533977303 | RP11-962G15.1 | G/C | 2.70 (1.89 - 3.84) | N/A | N/A | 4.51E-08 | N/A | 4.51E-08 | N/A |
| rs5987402 | N/A | G/T | 0.97 (0.96 - 0.99) | 0.98 (0.97 - 0.99) | N/A | 1.04E-04 | 8.60E-05 | 3.81E-08 | N/A |
| rs74653330 | OCA2 | C/T | 1.15 (1.10 - 1.19) | 1.17 (0.92 - 1.48) | N/A | 8.88E-13 | 0.21 | 3.90E-13 | N/A |
| rs75507031 | CASP8, LS2CR12 | C/T | 0.95 (0.94 - 0.97) | 0.92 (0.91 - 0.94) | N/A | 1.46E-08 | 2.07E-27 | 9.23E-33 | N/A |
| chr19:17104735 | N/A | A/ AAAAAAAC | N/A | 0.96 (0.95 - 0.98) | N/A | N/A | 2.95E-08 | 2.95E-08 | N/A |
| chr11:113707268 | N/A | A/ATTG | N/A | 1.05 (1.03 - 1.06) | N/A | N/A | 7.85E-09 | 7.85E-09 | N/A |
| chr5:1339775 | N/A | A/G | N/A | 0.95 (0.94 - 0.96) | N/A | N/A | 8.58E-15 | 8.58E-15 | N/A |

Supplementary Table S10. 62 Lead SNVs from FinnGen and UKB pan-cancer analysis and their overlap with the IPF meta-analysis.

### Contributors of FinnGen

#### Steering Committee

Aarno Palotie Institute for Molecular Medicine Finland, HiLIFE, University of Helsinki, Finland

Mark Daly Institute for Molecular Medicine Finland, HiLIFE, University of Helsinki, Finland

##### Pharmaceutical companies

Bridget Riley-Gills Abbvie, Chicago, IL, United States

Howard Jacob Abbvie, Chicago, IL, United States

Dirk Paul Astra Zeneca, Cambridge, United Kingdom

Heiko Runz Biogen, Cambridge, MA, United States

Sally John Biogen, Cambridge, MA, United States

Robert Plenge Celgene, Summit, NJ, United States/Bristol Myers Squibb, New York, NY, United States

Mark McCarthy Genentech, San Francisco, CA, United States

Julie Hunkapiller Genentech, San Francisco, CA, United States

Meg Ehm GlaxoSmithKline, Brentford, United Kingdom

Kirsi Auro GlaxoSmithKline, Brentford, United Kingdom

Caroline Fox Merck, Kenilworth, NJ, United States

Anders Mälarstig Pfizer, New York, NY, United States

Katherine Klinger Sanofi, Paris, France

Deepak Raipal Sanofi, Paris, France

Tim Behrens Maze Therapeutics, San Francisco, CA, United States

Robert Yang Janssen Biotech, Beerse, Belgium

Richard Siegel Novartis, Basel, Switzerland

##### University of Helsinki & Biobanks

Tomi Mäkelä HiLIFE, University of Helsinki, Finland, Finland

Jaakko Kaprio Institute for Molecular Medicine Finland, HiLIFE, Helsinki, Finland, Finland

Petri Virolainen Auria Biobank / University of Turku / Hospital District of Southwest Finland, Turku, Finland

Antti Hakanen Auria Biobank / University of Turku / Hospital District of Southwest Finland, Turku, Finland

Terhi Kilpi THL Biobank / The National Institute of Health and Welfare Helsinki, Finland

Markus Perola THL Biobank / The National Institute of Health and Welfare Helsinki, Finland

Jukka Partanen Finnish Red Cross Blood Service / Finnish Hematology Registry and Clinical Biobank, Helsinki, Finland

Anne Pitkäranta Helsinki Biobank / Helsinki University and Hospital District of Helsinki and Uusimaa, Helsinki

Juhani Junttila Northern Finland Biobank Borealis / University of Oulu / Northern Ostrobothnia Hospital District, Oulu, Finland

Raisa Serpi Northern Finland Biobank Borealis / University of Oulu / Northern Ostrobothnia Hospital District, Oulu, Finland

Tarja Laitinen Finnish Clinical Biobank Tampere / University of Tampere / Pirkanmaa Hospital District, Tampere, Finland

Johanna Mäkelä Finnish Clinical Biobank Tampere / University of Tampere / Pirkanmaa Hospital District, Tampere, Finland

Veli-Matti Kosma Biobank of Eastern Finland / University of Eastern Finland / Northern Savo Hospital District, Kuopio, Finland

Urho Kujala Central Finland Biobank / University of Jyväskylä / Central Finland Health Care District, Jyväskylä, Finland

**Other Experts/ Non-Voting Members**

Outi Tuovila Business Finland, Helsinki, Finland

Raimo Pakkanen Business Finland, Helsinki, Finland

#### Scientific Committee

**Pharmaceutical companies**

Jeffrey Waring Abbvie, Chicago, IL, United States

Ali Abbasi Abbvie, Chicago, IL, United States

Mengzhen Liu	Abbvie, Chicago, IL, United States

Ioanna Tachmazidou	Astra Zeneca, Cambridge, United Kingdom

Chia-Yen Chen	Biogen, Cambridge, MA, United States

Heiko Runz	Biogen, Cambridge, MA, United States

Shameek Biswas	Celgene, Summit, NJ, United States/Bristol Myers Squibb, New York, NY, United States

Julie Hunkapiller	Genentech, San Francisco, CA, United States

Meg Ehm	GlaxoSmithKline, Brentford, United Kingdom

Neha Raghavan	Merck, Kenilworth, NJ, United States

Adriana Huertas-Vazquez	Merck, Kenilworth, NJ, United States

Anders Mälarstig	Pfizer, New York, NY, United States

Xinli Hu	Pfizer, New York, NY, United States

Katherine Klinger	Sanofi, Paris, France

Matthias Gossel	Sanofi, Paris, France

Robert Graham	Maze Therapeutics, San Francisco, CA, United States

Tim Behrens	Maze Therapeutics, San Francisco, CA, United States

Beryl Cummings	Maze Therapeutics, San Francisco, CA, United States

Wilco Fleuren	Janssen Biotech, Beerse, Belgium

Dawn Waterworth	Janssen Biotech, Beerse, Belgium

Nicole Renaud	Novartis, Basel, Switzerland

Aviv Madar	Novartis, Basel, Switzerland

Maen Obeidat	Novartis, Basel, Switzerland

**University of Helsinki & Biobanks**

Samuli Ripatti	Institute for Molecular Medicine Finland, HiLIFE, Helsinki, Finland

Johanna Schleutker	Auria Biobank / Univ. of Turku / Hospital District of Southwest Finland, Turku, Finland

Markus Perola	THL Biobank / The National Institute of Health and Welfare Helsinki, Finland

Mikko Arvas Finnish Red Cross Blood Service / Finnish Hematology Registry and Clinical Biobank, Helsinki, Finland

Olli Carpén Helsinki Biobank / Helsinki University and Hospital District of Helsinki and Uusimaa, Helsinki

Reetta Hinttala Northern Finland Biobank Borealis / University of Oulu / Northern Ostrobothnia Hospital District, Oulu, Finland

Johannes Kettunen Northern Finland Biobank Borealis / University of Oulu / Northern Ostrobothnia Hospital District, Oulu, Finland

Johanna Mäkelä Finnish Clinical Biobank Tampere / University of Tampere / Pirkanmaa Hospital District, Tampere, Finland

Arto Mannermaa Biobank of Eastern Finland / University of Eastern Finland / Northern Savo Hospital District, Kuopio, Finland

Jari Laukkanen Central Finland Biobank / University of Jyväskylä / Central Finland Health Care District, Jyväskylä, Finland

Urho Kujala Central Finland Biobank / University of Jyväskylä / Central Finland Health Care District, Jyväskylä, Finland

#### Clinical Groups

**Neurology Group**

Reetta Kälviäinen Northern Savo Hospital District, Kuopio, Finland

Valtteri Julkunen Northern Savo Hospital District, Kuopio, Finland

Hilkka Soininen Northern Savo Hospital District, Kuopio, Finland

Anne Remes Northern Ostrobothnia Hospital District, Oulu, Finland

Mikko Hiltunen Northern Savo Hospital District, Kuopio, Finland

Jukka Peltola Pirkanmaa Hospital District, Tampere, Finland

Pentti Tienari Hospital District of Helsinki and Uusimaa, Helsinki, Finland

Juha Rinne Hospital District of Southwest Finland, Turku, Finland

Roosa Kallionpää Hospital District of Southwest Finland, Turku, Finland

Ali Abbasi Abbvie, Chicago, IL, United States

Adam Ziemann Abbvie, Chicago, IL, United States

Jeffrey Waring Abbvie, Chicago, IL, United States

Sahar Esmaeeli Abbvie, Chicago, IL, United States

Nizar Smaoui Abbvie, Chicago, IL, United States

Anne Lehtonen Abbvie, Chicago, IL, United States

Susan Eaton Biogen, Cambridge, MA, United States

Heiko Runz Biogen, Cambridge, MA, United States

Sanni Lahdenperä Biogen, Cambridge, MA, United States

Janet van Adelsberg Celgene, Summit, NJ, United States/ Bristol Myers Squibb, New York, NY, United States

Shameek Biswas Celgene, Summit, NJ, United States/ Bristol Myers Squibb, New York, NY, United States

Julie Hunkapiller Genentech, San Francisco, CA, United States

Natalie Bowers Genentech, San Francisco, CA, United States

Edmond Teng Genentech, San Francisco, CA, United States

Sarah Pendergrass Genentech, San Francisco, CA, United States

Onuralp Soylemez Merck, Kenilworth, NJ, United States

Kari Linden Pfizer, New York, NY, United States

Fanli Xu GlaxoSmithKline, Brentford, United Kingdom

David Pulford GlaxoSmithKline, Brentford, United Kingdom

Kirsi Auro GlaxoSmithKline, Brentford, United Kingdom

Laura Addis GlaxoSmithKline, Brentford, United Kingdom

John Eicher GlaxoSmithKline, Brentford, United Kingdom

Minna Raivio Hospital District of Helsinki and Uusimaa, Helsinki, Finland

Sarah Pendergrass Genentech, San Francisco, CA, United States

Beryl Cummings Maze Therapeutics, San Francisco, CA, United States

Juulia Partanen Institute for Molecular Medicine Finland, HiLIFE, University of Helsinki, Finland

**Gastroenterology Group**

Martti Färkkilä Hospital District of Helsinki and Uusimaa, Helsinki, Finland

Jukka Koskela Hospital District of Helsinki and Uusimaa, Helsinki, Finland

Sampsa Pikkarainen Hospital District of Helsinki and Uusimaa, Helsinki, Finland

Airi Jussila Pirkanmaa Hospital District, Tampere, Finland

Katri Kaukinen Pirkanmaa Hospital District, Tampere, Finland

Timo Blomster Northern Ostrobothnia Hospital District, Oulu, Finland

Mikko Kiviniemi Northern Savo Hospital District, Kuopio, Finland

Markku Voutilainen Hospital District of Southwest Finland, Turku, Finland

Ali Abbasi Abbvie, Chicago, IL, United States

Graham Heap Abbvie, Chicago, IL, United States

Jeffrey Waring Abbvie, Chicago, IL, United States

Nizar Smaoui Abbvie, Chicago, IL, United States

Fedik Rahimov Abbvie, Chicago, IL, United States

Anne Lehtonen Abbvie, Chicago, IL, United States

Keith Usiskin Celgene, Summit, NJ, United States/ Bristol Myers Squibb, New York, NY, United States

Tim Lu Genentech, San Francisco, CA, United States

Natalie Bowers Genentech, San Francisco, CA, United States

Danny Oh Genentech, San Francisco, CA, United States

Sarah Pendergrass Genentech, San Francisco, CA, United States

Kirsi Kalpala Pfizer, New York, NY, United States

Melissa Miller Pfizer, New York, NY, United States

Xinli Hu Pfizer, New York, NY, United States

Linda McCarthy GlaxoSmithKline, Brentford, United Kingdom

Onuralp Soylemez Merck, Kenilworth, NJ, United States

Mark Daly Institute for Molecular Medicine Finland, HiLIFE, University of Helsinki, Finland

**Rheumatology Group**

Kari Eklund Hospital District of Helsinki and Uusimaa, Helsinki, Finland

Antti Palomäki Hospital District of Southwest Finland, Turku, Finland

Pia Isomäki Pirkanmaa Hospital District, Tampere, Finland

Laura Pirilä Hospital District of Southwest Finland, Turku, Finland

Oili Kaipiainen-Seppänen Northern Savo Hospital District, Kuopio, Finland

Johanna Huhtakangas Northern Ostrobothnia Hospital District, Oulu, Finland

Ali Abbasi Abbvie, Chicago, IL, United States

Jeffrey Waring Abbvie, Chicago, IL, United States

Fedik Rahimov Abbvie, Chicago, IL, United States

Apinya Lertratanakul Abbvie, Chicago, IL, United States

Nizar Smaoui Abbvie, Chicago, IL, United States

Anne Lehtonen Abbvie, Chicago, IL, United States

David Close Astra Zeneca, Cambridge, United Kingdom

Marla Hochfeld Celgene, Summit, NJ, United States/ Bristol Myers Squibb, New York, NY, United States

Natalie Bowers Genentech, San Francisco, CA, United States

Sarah Pendergrass Genentech, San Francisco, CA, United States

Onuralp Soylemez Merck, Kenilworth, NJ, United States

Kirsi Kalpala Pfizer, New York, NY, United States

Nan Bing Pfizer, New York, NY, United States

Xinli Hu Pfizer, New York, NY, United States

Jorge Esparza Gordillo GlaxoSmithKline, Brentford, United Kingdom

Kirsi Auro GlaxoSmithKline, Brentford, United Kingdom

Dawn Waterworth Janssen Biotech, Beerse, Belgium

Nina Mars Institute for Molecular Medicine Finland, HiLIFE, Helsinki, Finland

**Pulmonology Group**

Tarja Laitinen Pirkanmaa Hospital District, Tampere, Finland

Margit Pelkonen Northern Savo Hospital District, Kuopio, Finland

Paula Kauppi Hospital District of Helsinki and Uusimaa, Helsinki, Finland

Hannu Kankaanranta Pirkanmaa Hospital District, Tampere, Finland

Terttu Harju Northern Ostrobothnia Hospital District, Oulu, Finland

Riitta Lahesmaa Hospital District of Southwest Finland, Turku, Finland

Nizar Smaoui Abbvie, Chicago, IL, United States

Alex Mackay Astra Zeneca, Cambridge, United Kingdom

Glenda Lassi Astra Zeneca, Cambridge, United Kingdom

Susan Eaton Biogen, Cambridge, MA, United States

Steven Greenberg Celgene, Summit, NJ, United States/ Bristol Myers Squibb, New York, NY, United States

Hubert Chen Genentech, San Francisco, CA, United States

Sarah Pendergrass Genentech, San Francisco, CA, United States

Natalie Bowers Genentech, San Francisco, CA, United States

Joanna Betts GlaxoSmithKline, Brentford, United Kingdom

Soumitra Ghosh GlaxoSmithKline, Brentford, United Kingdom

Kirsi Auro GlaxoSmithKline, Brentford, United Kingdom

Rajashree Mishra GlaxoSmithKline, Brentford, United Kingdom

Sina Rüeger Institute for Molecular Medicine Finland, HiLIFE, University of Helsinki, Finland

**Cardiometabolic Diseases Group**

Teemu Niiranen The National Institute of Health and Welfare Helsinki, Finland

Felix Vaura The National Institute of Health and Welfare Helsinki, Finland

Veikko Salomaa The National Institute of Health and Welfare Helsinki, Finland

Markus Juonala Hospital District of Southwest Finland, Turku, Finland

Kaj Metsärinne Hospital District of Southwest Finland, Turku, Finland

Mika Kähönen Pirkanmaa Hospital District, Tampere, Finland

Juhani Junttila Northern Ostrobothnia Hospital District, Oulu, Finland

Markku Laakso Northern Savo Hospital District, Kuopio, Finland

Jussi Pihlajamäki Northern Savo Hospital District, Kuopio, Finland

Daniel Gordin Hospital District of Helsinki and Uusimaa, Helsinki, Finland

Juha Sinisalo Hospital District of Helsinki and Uusimaa, Helsinki, Finland

Marja-Riitta Taskinen Hospital District of Helsinki and Uusimaa, Helsinki, Finland

Tiinamaija Tuomi Hospital District of Helsinki and Uusimaa, Helsinki, Finland

Jari Laukkanen Central Finland Health Care District, Jyväskylä, Finland

Benjamin Challis Astra Zeneca, Cambridge, United Kingdom

Dirk Paul Astra Zeneca, Cambridge, United Kingdom

Julie Hunkapiller Genentech, San Francisco, CA, United States

Natalie Bowers Genentech, San Francisco, CA, United States

Sarah Pendergrass Genentech, San Francisco, CA, United States

Onuralp Soylemez Merck, Kenilworth, NJ, United States

Jaakko Parkkinen Pfizer, New York, NY, United States

Melissa Miller Pfizer, New York, NY, United States

Russell Miller Pfizer, New York, NY, United States

Audrey Chu GlaxoSmithKline, Brentford, United Kingdom

Kirsi Auro GlaxoSmithKline, Brentford, United Kingdom

Keith Usiskin Celgene, Summit, NJ, United States/ Bristol Myers Squibb, New York, NY, United States

Amanda Elliott Institute for Molecular Medicine Finland, HiLIFE, University of Helsinki, Finland / Broad Institute, Cambridge, MA, United States

Joel Rämö Institute for Molecular Medicine Finland, HiLIFE, University of Helsinki, Finland

Samuli Ripatti Institute for Molecular Medicine Finland, HiLIFE, University of Helsinki, Finland

Mary Pat Reeve Institute for Molecular Medicine Finland, HiLIFE, University of Helsinki, Finland

Sanni Ruotsalainen Institute for Molecular Medicine Finland, HiLIFE, University of Helsinki, Finland

**Oncology Group**

Tuomo Meretoja Hospital District of Helsinki and Uusimaa, Helsinki, Finland

Heikki Joensuu Hospital District of Helsinki and Uusimaa, Helsinki, Finland

Olli Carpén Hospital District of Helsinki and Uusimaa, Helsinki, Finland

Lauri Aaltonen Hospital District of Helsinki and Uusimaa, Helsinki, Finland

Johanna Mattson Hospital District of Helsinki and Uusimaa, Helsinki, Finland

Annika Auranen Pirkanmaa Hospital District , Tampere, Finland

| | |
|---|---|
| Peeter Karihtala | Northern Ostrobothnia Hospital District, Oulu, Finland |
| Saila Kauppila | Northern Ostrobothnia Hospital District, Oulu, Finland |
| Päivi Auvinen | Northern Savo Hospital District, Kuopio, Finland |
| Klaus Elenius | Hospital District of Southwest Finland, Turku, Finland |
| Johanna Schleutker | Hospital District of Southwest Finland, Turku, Finland |
| Relja Popovic | Abbvie, Chicago, IL, United States |
| Jeffrey Waring | Abbvie, Chicago, IL, United States |
| Bridget Riley-Gillis | Abbvie, Chicago, IL, United States |
| Anne Lehtonen | Abbvie, Chicago, IL, United States |
| Jennifer Schutzman | Genentech, San Francisco, CA, United States |
| Julie Hunkapiller | Genentech, San Francisco, CA, United States |
| Natalie Bowers | Genentech, San Francisco, CA, United States |
| Sarah Pendergrass | Genentech, San Francisco, CA, United States |
| Andrey Loboda | Merck, Kenilworth, NJ, United States |
| Aparna Chhibber | Merck, Kenilworth, NJ, United States |
| Heli Lehtonen | Pfizer, New York, NY, United States |
| Stefan McDonough | Pfizer, New York, NY, United States |
| Marika Crohns | Sanofi, Paris, France |
| Sauli Vuoti | Sanofi, Paris, France |
| Diptee Kulkarni | GlaxoSmithKline, Brentford, United Kingdom |
| Kirsi Auro | GlaxoSmithKline, Brentford, United Kingdom |
| Esa Pitkänen | Institute for Molecular Medicine Finland, HiLIFE, University of Helsinki, Finland |
| Nina Mars | Institute for Molecular Medicine Finland, HiLIFE, University of Helsinki, Finland |
| Mark Daly | Institute for Molecular Medicine Finland, HiLIFE, University of Helsinki, Finland |

**Opthalmology Group**

| | |
|---|---|
| Kai Kaarniranta | Northern Savo Hospital District, Kuopio, Finland |
| Joni A Turunen | Hospital District of Helsinki and Uusimaa, Helsinki, Finland |
| Terhi Ollila | Hospital District of Helsinki and Uusimaa, Helsinki, Finland |
| Sanna Seitsonen | Hospital District of Helsinki and Uusimaa, Helsinki, Finland |

Hannu Uusitalo Pirkanmaa Hospital District, Tampere, Finland

Vesa Aaltonen Hospital District of Southwest Finland, Turku, Finland

Hannele Uusitalo-Järvinen Pirkanmaa Hospital District, Tampere, Finland

Marja Luodonpää Northern Ostrobothnia Hospital District, Oulu, Finland

Nina Hautala Northern Ostrobothnia Hospital District, Oulu, Finland

Mengzhen Liu Abbvie, Chicago, IL, United States

Heiko Runz Biogen, Cambridge, MA, United States

Stephanie Loomis Biogen, Cambridge, MA, United States

Erich Strauss Genentech, San Francisco, CA, United States

Natalie Bowers Genentech, San Francisco, CA, United States

Hao Chen Genentech, San Francisco, CA, United States

Sarah Pendergrass Genentech, San Francisco, CA, United States

Anna Podgornaia Merck, Kenilworth, NJ, United States

Juha Karjalainen Institute for Molecular Medicine Finland, HiLIFE, University of Helsinki, Finland / Broad Institute, Cambridge, MA, United States

Esa Pitkänen Institute for Molecular Medicine Finland, HiLIFE, University of Helsinki, Finland

**Dermatology Group**

Kaisa Tasanen Northern Ostrobothnia Hospital District, Oulu, Finland

Laura Huilaja Northern Ostrobothnia Hospital District, Oulu, Finland

Katariina Hannula-Jouppi Hospital District of Helsinki and Uusimaa, Helsinki, Finland

Teea Salmi Pirkanmaa Hospital District, Tampere, Finland

Sirkku Peltonen Hospital District of Southwest Finland, Turku, Finland

Leena Koulu Hospital District of Southwest Finland, Turku, Finland

Kirsi Kalpala Pfizer, New York, NY, United States

Ying Wu Pfizer, New York, NY, United States

David Choy Genentech, San Francisco, CA, United States

Sarah Pendergrass Genentech, San Francisco, CA, United States

Nizar Smaoui Abbvie, Chicago, IL, United States

Fedik Rahimov Abbvie, Chicago, IL, United States

| | |
|---|---|
| Anne Lehtonen | Abbvie, Chicago, IL, United States |
| Dawn Waterworth | Janssen Biotech, Beerse, Belgium |

**Odontology Group**

| | |
|---|---|
| Pirkko Pussinen | Hospital District of Helsinki and Uusimaa, Helsinki, Finland |
| Aino Salminen | Hospital District of Helsinki and Uusimaa, Helsinki, Finland |
| Tuula Salo | Hospital District of Helsinki and Uusimaa, Helsinki, Finland |
| David Rice | Hospital District of Helsinki and Uusimaa, Helsinki, Finland |
| Pekka Nieminen | Hospital District of Helsinki and Uusimaa, Helsinki, Finland |
| Ulla Palotie | Hospital District of Helsinki and Uusimaa, Helsinki, Finland |
| Juha Sinisalo | Hospital District of Helsinki and Uusimaa, Helsinki, Finland |
| Maria Siponen | Northern Savo Hospital District, Kuopio, Finland |
| Liisa Suominen | Northern Savo Hospital District, Kuopio, Finland |
| Päivi Mäntylä | Northern Savo Hospital District, Kuopio, Finland |
| Ulvi Gursoy | Hospital District of Southwest Finland, Turku, Finland |
| Vuokko Anttonen | Northern Ostrobothnia Hospital District, Oulu, Finland |
| Kirsi Sipilä | Northern Ostrobothnia Hospital District, Oulu, Finland |
| Sarah Pendergrass | Genentech, San Francisco, CA, United States |

**Women's Health and Reproduction Group**

| | |
|---|---|
| Hannele Laivuori | Institute for Molecular Medicine Finland, HiLIFE, University of Helsinki, Finland |
| Venla Kurra | Pirkanmaa Hospital District, Tampere, Finland |
| Oskari Heikinheimo | Hospital District of Helsinki and Uusimaa, Helsinki, Finland |
| Ilkka Kalliala | Hospital District of Helsinki and Uusimaa, Helsinki, Finland |
| Laura Kotaniemi-Talonen | Pirkanmaa Hospital District, Tampere, Finland |
| Kari Nieminen | Pirkanmaa Hospital District, Tampere, Finland |
| Päivi Polo | Hospital District of Southwest Finland, Turku, Finland |
| Kaarin Mäkikallio | Hospital District of Southwest Finland, Turku, Finland |
| Eeva Ekholm | Hospital District of Southwest Finland, Turku, Finland |
| Marja Vääräsmäki | Northern Ostrobothnia Hospital District, Oulu, Finland |

Outi Uimari Northern Ostrobothnia Hospital District, Oulu, Finland

Laure Morin-Papunen Northern Ostrobothnia Hospital District, Oulu, Finland

Marjo Tuppurainen Northern Savo Hospital District, Kuopio, Finland

Katja Kivinen Institute for Molecular Medicine Finland, HiLIFE, University of Helsinki, Finland

Elisabeth Widen Institute for Molecular Medicine Finland, HiLIFE, University of Helsinki, Finland

Taru Tukiainen Institute for Molecular Medicine Finland, HiLIFE, University of Helsinki, Finland

Mary Pat Reeve Institute for Molecular Medicine Finland, HiLIFE, University of Helsinki, Finland

Mark Daly Institute for Molecular Medicine Finland, HiLIFE, University of Helsinki, Finland

Liu Aoxing Institute for Molecular Medicine Finland, HiLIFE, University of Helsinki, Finland

Eija Laakkonen University of Jyväskylä, Jyväskylä, Finland

Niko Välimäki University of Helsinki, Helsinki, Finland

Lauri Aaltonen Hospital District of Helsinki and Uusimaa, Helsinki, Finland

Johannes Kettunen Northern Ostrobothnia Hospital District, Oulu, Finland

Mikko Arvas Finnish Red Cross Blood Service, Helsinki, Finland

Jeffrey Waring Abbvie, Chicago, IL, United States

Bridget Riley-Gillis Abbvie, Chicago, IL, United States

Mengzhen Liu Abbvie, Chicago, IL, United States

Janet Kumar GlaxoSmithKline, Brentford, United Kingdom

Kirsi Auro GlaxoSmithKline, Brentford, United Kingdom

Andrea Ganna Institute for Molecular Medicine Finland, HiLIFE, University of Helsinki, Finland

Sarah Pendergrass Genentech, San Francisco, CA, United States

##### FinnGen Analysis working group

Justin Wade Davis Abbvie, Chicago, IL, United States

Bridget Riley-Gillis Abbvie, Chicago, IL, United States

Danjuma Quarless Abbvie, Chicago, IL, United States

Fedik Rahimov Abbvie, Chicago, IL, United States

Sahar Esmaeeli Abbvie, Chicago, IL, United States

Slavé Petrovski Astra Zeneca, Cambridge, United Kingdom

Eleonor Wigmore Astra Zeneca, Cambridge, United Kingdom

Adele Mitchell Biogen, Cambridge, MA, United States

Benjamin Sun Biogen, Cambridge, MA, United States

Ellen Tsai Biogen, Cambridge, MA, United States

Denis Baird Biogen, Cambridge, MA, United States

Paola Bronson Biogen, Cambridge, MA, United States

Ruoyu Tian Biogen, Cambridge, MA, United States

Stephanie Loomis Biogen, Cambridge, MA, United States

Yunfeng Huang Biogen, Cambridge, MA, United States

Joseph Maranville Celgene, Summit, NJ, United States/ Bristol Myers Squibb, New York, NY, United States

Shameek Biswas Celgene, Summit, NJ, United States/ Bristol Myers Squibb, New York, NY, United States

Elmutaz Mohammed Celgene, Summit, NJ, United States/ Bristol Myers Squibb, New York, NY, United States

Samir Wadhawan Celgene, Summit, NJ, United States/ Bristol Myers Squibb, New York, NY, United States

Erika Kvikstad Celgene, Summit, NJ, United States/ Bristol Myers Squibb, New York, NY, United States

Minal Caliskan Celgene, Summit, NJ, United States/ Bristol Myers Squibb, New York, NY, United States

Diana Chang Genentech, San Francisco, CA, United States

Julie Hunkapiller Genentech, San Francisco, CA, United States

Tushar Bhangale Genentech, San Francisco, CA, United States

Natalie Bowers Genentech, San Francisco, CA, United States

Sarah Pendergrass Genentech, San Francisco, CA, United States

Kirill Shkura Merck, Kenilworth, NJ, United States

Victor Neduva Merck, Kenilworth, NJ, United States

Xing Chen Pfizer, New York, NY, United States

Åsa Hedman Pfizer, New York, NY, United States

Karen S King GlaxoSmithKline, Brentford, United Kingdom

Padhraig Gormley GlaxoSmithKline, Brentford, United Kingdom

Jimmy Liu GlaxoSmithKline, Brentford, United Kingdom

Clarence Wang Sanofi, Paris, France

Ethan Xu Sanofi, Paris, France

Franck Auge Sanofi, Paris, France

Clement Chatelain Sanofi, Paris, France

Deepak Rajpal Sanofi, Paris, France

Dongyu Liu Sanofi, Paris, France

Katherine Call Sanofi, Paris, France

Tai-He Xia Sanofi, Paris, France

Beryl Cummings Maze Therapeutics, San Francisco, CA, United States

Matt Brauer Maze Therapeutics, San Francisco, CA, United States

Huilei Xu Novartis, Basel, Switzerland

Amy Cole Novartis, Basel, Switzerland

Jonathan Chung Novartis, Basel, Switzerland

Jaison Jacob Novartis, Basel, Switzerland

Katrina de Lange Novartis, Basel, Switzerland

Jonas Zierer Novartis, Basel, Switzerland

Mitja Kurki Institute for Molecular Medicine Finland, HiLIFE, University of Helsinki, Finland / Broad Institute, Cambridge, MA, United States

Samuli Ripatti Institute for Molecular Medicine Finland, HiLIFE, University of Helsinki, Finland

Mark Daly Institute for Molecular Medicine Finland, HiLIFE, University of Helsinki, Finland

Juha Karjalainen Institute for Molecular Medicine Finland, HiLIFE, University of Helsinki, Finland / Broad Institute, Cambridge, MA, United States

Aki Havulinna Institute for Molecular Medicine Finland, HiLIFE, University of Helsinki, Finland

Juha Mehtonen Institute for Molecular Medicine Finland, HiLIFE, University of Helsinki, Finland

Priit Palta Institute for Molecular Medicine Finland, HiLIFE, University of Helsinki, Finland

Shabbeer Hassan Institute for Molecular Medicine Finland, HiLIFE, University of Helsinki, Finland

Pietro Della Briotta Parolo Institute for Molecular Medicine Finland, HiLIFE, University of Helsinki, Finland

Wei Zhou Broad Institute, Cambridge, MA, United States

Mutaamba Maasha Broad Institute, Cambridge, MA, United States

Shabbeer Hassan Institute for Molecular Medicine Finland, HiLIFE, University of Helsinki, Finland

Susanna Lemmelä Institute for Molecular Medicine Finland, HiLIFE, University of Helsinki, Finland

Manuel Rivas University of Stanford, Stanford, CA, United States

Aarno Palotie Institute for Molecular Medicine Finland, HiLIFE, University of Helsinki, Finland

Arto Lehisto Institute for Molecular Medicine Finland, HiLIFE, University of Helsinki, Finland

Andrea Ganna Institute for Molecular Medicine Finland, HiLIFE, University of Helsinki, Finland

Vincent Llorens Institute for Molecular Medicine Finland, HiLIFE, University of Helsinki, Finland

Hannele Laivuori Institute for Molecular Medicine Finland, HiLIFE, University of Helsinki, Finland

Mari E Niemi Institute for Molecular Medicine Finland, HiLIFE, University of Helsinki, Finland

Taru Tukiainen Institute for Molecular Medicine Finland, HiLIFE, University of Helsinki, Finland

Mary Pat Reeve Institute for Molecular Medicine Finland, HiLIFE, University of Helsinki, Finland

Henrike Heyne Institute for Molecular Medicine Finland, HiLIFE, University of Helsinki, Finland

Nina Mars Institute for Molecular Medicine Finland, HiLIFE, University of Helsinki, Finland

Kimmo Palin University of Helsinki, Helsinki, Finland

Javier Garcia-Tabuenca University of Tampere, Tampere, Finland

Harri Siirtola University of Tampere, Tampere, Finland

Tuomo Kiiskinen Institute for Molecular Medicine Finland, HiLIFE, University of Helsinki, Finland

Jiwoo Lee Institute for Molecular Medicine Finland, HiLIFE, University of Helsinki, Finland / Broad Institute, Cambridge, MA, United States

Kristin Tsuo Institute for Molecular Medicine Finland, HiLIFE, University of Helsinki, Finland / Broad Institute, Cambridge, MA, United States

Amanda Elliott Institute for Molecular Medicine Finland, HiLIFE, University of Helsinki, Finland / Broad Institute, Cambridge, MA, United States

Kati Kristiansson THL Biobank / The National Institute of Health and Welfare Helsinki, Finland

Mikko Arvas Finnish Red Cross Blood Service / Finnish Hematology Registry and Clinical Biobank, Helsinki, Finland

Kati Hyvärinen Finnish Red Cross Blood Service, Helsinki, Finland

Jarmo Ritari Finnish Red Cross Blood Service, Helsinki, Finland

Miika Koskinen Helsinki Biobank / Helsinki University and Hospital District of Helsinki and Uusimaa, Helsinki

Olli Carpén Helsinki Biobank / Helsinki University and Hospital District of Helsinki and Uusimaa, Helsinki

Johannes Kettunen Northern Finland Biobank Borealis / University of Oulu / Northern Ostrobothnia Hospital District, Oulu, Finland

Katri Pylkäs University of Oulu, Oulu, Finland

Marita Kalaoja University of Oulu, Oulu, Finland

Minna Karjalainen University of Oulu, Oulu, Finland

Tuomo Mantere Northern Finland Biobank Borealis / University of Oulu / Northern Ostrobothnia Hospital District, Oulu, Finland

Eeva Kangasniemi Finnish Clinical Biobank Tampere / University of Tampere / Pirkanmaa Hospital District, Tampere, Finland

Sami Heikkinen University of Eastern Finland, Kuopio, Finland

Arto Mannermaa Biobank of Eastern Finland / University of Eastern Finland / Northern Savo Hospital District, Kuopio, Finland

Eija Laakkonen University of Jyväskylä, Jyväskylä, Finland

Samuel Heron University of Turku, Turku, Finland

Dhanaprakash Jambulingam University of Turku, Turku, Finland

Venkat Subramaniam Rathinakannan University of Turku, Turku, Finland

Nina Pitkänen Auria Biobank / University of Turku / Hospital District of Southwest Finland, Turku, Finland

#### Biobank directors

Lila Kallio Auria Biobank / University of Turku / Hospital District of Southwest Finland, Turku, Finland

Sirpa Soini THL Biobank / The National Institute of Health and Welfare Helsinki, Finland

Jukka Partanen Finnish Red Cross Blood Service / Finnish Hematology Registry and Clinical Biobank, Helsinki, Finland

Eero Punkka Helsinki Biobank / Helsinki University and Hospital District of Helsinki and Uusimaa, Helsinki

Raisa Serpi Northern Finland Biobank Borealis / University of Oulu / Northern Ostrobothnia Hospital District, Oulu, Finland

Johanna Mäkelä Finnish Clinical Biobank Tampere / University of Tampere / Pirkanmaa Hospital District, Tampere, Finland

Veli-Matti Kosma Biobank of Eastern Finland / University of Eastern Finland / Northern Savo Hospital District, Kuopio, Finland

Teijo Kuopio Central Finland Biobank / University of Jyväskylä / Central Finland Health Care District, Jyväskylä, Finland

#### FinnGen Teams

##### Administration

Anu Jalanko Institute for Molecular Medicine Finland, HiLIFE, University of Helsinki, Finland

Huei-Yi Shen Institute for Molecular Medicine Finland, HiLIFE, University of Helsinki, Finland

Risto Kajanne Institute for Molecular Medicine Finland, HiLIFE, University of Helsinki, Finland

Mervi Aavikko Institute for Molecular Medicine Finland, HiLIFE, University of Helsinki, Finland

##### Analysis

Mitja Kurki Institute for Molecular Medicine Finland, HiLIFE, University of Helsinki, Finland / Broad Institute, Cambridge, MA, United States

Juha Karjalainen Institute for Molecular Medicine Finland, HiLIFE, University of Helsinki, Finland / Broad Institute, Cambridge, MA, United States

Pietro Della Briotta Parolo Institute for Molecular Medicine Finland, HiLIFE, University of Helsinki, Finland

Arto Lehisto Institute for Molecular Medicine Finland, HiLIFE, University of Helsinki, Finland

Juha Mehtonen Institute for Molecular Medicine Finland, HiLIFE, University of Helsinki, Finland

Wei Zhou Broad Institute, Cambridge, MA, United States

Masahiro Kanai Broad Institute, Cambridge, MA, United States

Mutaamba Maasha Broad Institute, Cambridge, MA, United States

**Clinical Endpoint Development**

Hannele Laivuori Institute for Molecular Medicine Finland, HiLIFE, University of Helsinki, Finland

Aki Havulinna Institute for Molecular Medicine Finland, HiLIFE, University of Helsinki, Finland

Susanna Lemmelä Institute for Molecular Medicine Finland, HiLIFE, University of Helsinki, Finland

Tuomo Kiiskinen Institute for Molecular Medicine Finland, HiLIFE, University of Helsinki, Finland

L. Elisa Lahtela Institute for Molecular Medicine Finland, HiLIFE, University of Helsinki, Finland

Matti Peura Institute for Molecular Medicine Finland, HiLIFE, University of Helsinki, Finland

**Communication**

Mari Kaunisto Institute for Molecular Medicine Finland, HiLIFE, University of Helsinki, Finland

**Data Management and IT Infrastructure**

Elina Kilpeläinen Institute for Molecular Medicine Finland, HiLIFE, University of Helsinki, Finland

Timo P. Sipilä Institute for Molecular Medicine Finland, HiLIFE, University of Helsinki, Finland

Georg Brein Institute for Molecular Medicine Finland, HiLIFE, University of Helsinki, Finland

Oluwaseun A. Dada Institute for Molecular Medicine Finland, HiLIFE, University of Helsinki, Finland

Awaisa Ghazal Institute for Molecular Medicine Finland, HiLIFE, University of Helsinki, Finland

Anastasia Shcherban Institute for Molecular Medicine Finland, HiLIFE, University of Helsinki, Finland

**Genotyping**

Kati Donner Institute for Molecular Medicine Finland, HiLIFE, University of Helsinki, Finland

Timo P. Sipilä Institute for Molecular Medicine Finland, HiLIFE, University of Helsinki, Finland

**Sample Collection Coordination**

Anu Loukola Helsinki Biobank / Helsinki University and Hospital District of Helsinki and Uusimaa, Helsinki

**Sample Logistics**

Päivi Laiho THL Biobank / The National Institute of Health and Welfare Helsinki, Finland

Tuuli Sistonen THL Biobank / The National Institute of Health and Welfare Helsinki, Finland

Essi Kaiharju THL Biobank / The National Institute of Health and Welfare Helsinki, Finland

Markku Laukkanen THL Biobank / The National Institute of Health and Welfare Helsinki, Finland

Elina Järvensivu THL Biobank / The National Institute of Health and Welfare Helsinki, Finland

Sini Lähteenmäki THL Biobank / The National Institute of Health and Welfare Helsinki, Finland

Lotta Männikkö THL Biobank / The National Institute of Health and Welfare Helsinki, Finland

Regis Wong THL Biobank / The National Institute of Health and Welfare Helsinki, Finland

**Registry Data Operations**

Hannele Mattsson THL Biobank / The National Institute of Health and Welfare Helsinki, Finland

Kati Kristiansson THL Biobank / The National Institute of Health and Welfare Helsinki, Finland

Susanna Lemmelä Institute for Molecular Medicine Finland, HiLIFE, University of Helsinki, Finland

Sami Koskelainen THL Biobank / The National Institute of Health and Welfare Helsinki, Finland

Tero Hiekkalinna THL Biobank / The National Institute of Health and Welfare Helsinki, Finland

Teemu Paajanen THL Biobank / The National Institute of Health and Welfare Helsinki, Finland

**Sequencing Informatics**

Priit Palta Institute for Molecular Medicine Finland, HiLIFE, University of Helsinki, Finland

Kalle Pärn Institute for Molecular Medicine Finland, HiLIFE, University of Helsinki, Finland

Shuang Luo Institute for Molecular Medicine Finland, HiLIFE, University of Helsinki, Finland

Vishal Sinha Institute for Molecular Medicine Finland, HiLIFE, University of Helsinki, Finland

**Trajectory Team**

Tarja Laitinen Pirkanmaa Hospital District, Tampere, Finland

Harri Siirtola University of Tampere, Tampere, Finland

Javier Gracia-Tabuenca University of Tampere, Tampere, Finland

Mika Helminen University of Tampere, Tampere, Finland

Tiina Luukkaala University of Tampere, Tampere, Finland

Iida Vähätalo University of Tampere, Tampere, Finland

**Data protection officer**

Tero Jyrhämä Institute for Molecular Medicine Finland, HiLIFE, University of Helsinki, Finland

**FinBB - Finnish biobank cooperative**

Marco Hautalahti

Laura Mustaniemi

Mirkka Koivusalo

Sarah Smith

Tom Southerington
